# Decoding Non-coding SNPs: Systems Genomics Modelling Dissects the Heterogeneity of IBD

**DOI:** 10.1101/2024.10.28.24316194

**Authors:** Dezső Módos, John P. Thomas, Johanne Brooks-Warburton, Martina Poletti, Balazs Bohar, Matthew Madgwick, Luca Csabai, Wen-Xin Kang, Benjamin Alexander-Dann, Azedine Zoufir, Padhmanand Sudhakar, Domenico Cozzetto, David Fazekas, Simon R. Carding, Nicholas Powell, Bram Verstockt, Andreas Bender, Tamas Korcsmaros

## Abstract

Genome-wide association studies have pinpointed numerous susceptibility loci in complex diseases like chronic immune-mediated inflammatory disorders (IMIDs), yet their impact on pathomechanisms remain poorly understood. Genetic epistasis, low effect sizes, and predominance within non-coding genomic regions, remain major challenges to the functional interpretation of IMID-associated single nucleotide polymorphisms (SNPs). To address this, we present a novel systems genomics approach which models the cumulative impact of non-coding SNPs on downstream cellular signalling and gene regulatory networks (GRNs). Applying this to the prototypical chronic IMIDs of Crohn’s disease (CD) and ulcerative colitis (UC), both forms of inflammatory bowel disease (IBD), we individually analysed 2,636 patient genomes. Signals from non-coding SNPs were found to propagate towards well-established and novel CD- and UC-associated pathogenic pathways through the signalling and gene regulatory layers. The SNP-propagated GRNs stratified CD and UC patients into distinct clusters corresponding to cell type-specific gene dysregulation and therapeutic response. This approach bridges the gap between genotype and phenotype, laying the foundations for accelerating precision medicine in complex diseases.

## Introduction

Following the sequencing of the human genome at the turn of the 21st century, genome-wide association studies (GWAS) have endeavoured to identify the genomic loci underpinning the heritability of common, complex traits and diseases^1^. These efforts have identified hundreds of susceptibility loci in chronic immune-mediated inflammatory disorders (IMIDs) such as rheumatoid arthritis and inflammatory bowel diseases (IBD) ^2^. Despite initial hopes that this would translate into a deeper understanding of disease pathogenesis and facilitate precision medicine strategies, this has yet to materialise ^3^. Several factors have contributed to this shortcoming. Firstly, the vast majority of risk loci in chronic IMIDs map to non-coding regions of the genome which are challenging to functionally analyse and interpret ^4,5^. Furthermore, individual risk variants exert small effect sizes and in combination explain only a small proportion of the total heritability ^6^, resulting in significant “missing heritability” ^7,8^. These have proven to be major obstacles for unravelling the genetic basis of complex diseases.

Accumulating evidence suggests that this missing heritability can in part be explained by the impact of non-coding genetic variants on gene regulation - in particular, the expression of *cis*-linked genes ^9,10^. This phenomenon has recently been coined as “missing regulation” ^11^. Another major factor contributing to missing heritability, and indeed, missing regulation is genetic epistasis i.e. multiple susceptibility loci can interact with each other, resulting in small perturbations in interconnected biological pathways which associate with the disease phenotype ^12^. These challenges mean that traditional approaches that interrogate the impact of individual susceptibility loci on phenotype in cell culture or animal models may not be suitable or feasible for unravelling genotype-to-phenotype associations in complex genetic diseases ^13,14^. Thus, there is a need for more sophisticated analytical models that study the collective impact of multiple non-coding variants on gene regulation, gene expression, and, ultimately, phenotype in complex diseases ^11^.

Systems genomics has recently emerged as a powerful approach that has the potential to address this specific need. Systems genomics utilises network biology principles to holistically detect the salient molecular interactions occurring within complex biological systems to identify gene-to-disease associations. Initial systems genomics methods focused on identifying the direct interactors and local neighbourhoods of seed genes within biological networks to infer biological pathways or functions associated with a disease ^15^. This is based on the guilt-by-association principle that genes or proteins that are closely related to each other due to co-expression or physical interactions are more likely to be involved in the same biological processes or pathways ^16^. We previously generated a guilt-by-association-based systems genomics workflow: the integrated Single Nucleotide Polymorphism (iSNP) network pipeline ^17^. Using patient genotype data, we predicted the combinatory effects of non-coding SNPs located within miRNA target sites and transcription factor binding sites (TFBS) in promoter and enhancer regions by identifying SNP-affected proteins and their first neighbour interactors in a signalling network. Whilst such local neighbourhood methods can provide useful biological insights, they do not capture the effect of genes or proteins on more distant connections and thus fail to fully contextualise seed genes or proteins within the global topology of biological networks ^18,19^. To address this limitation, novel methods have emerged based on the paradigm of network propagation that can amplify biological signals across the global architecture of biological networks ^16^. In network propagation, seed genes or proteins are considered as “hot spots” within the network and their “heat” diffuses to identify other “hot” candidates across the entire network through an iterative process ^16,18^. Importantly, network propagation has been found to outperform local neighbourhood methods to infer gene-to-disease associations, largely from studies in the field of oncology ^16^. Recently, random walk-based network propagation was employed to expand the known network of pleiotropic GWAS signals across multiple human traits, including chronic IMIDs, to reveal novel candidate genes and drug targets ^20^. However, there is a critical need to test the real-world applicability of such systems genomics approaches in adequately powered patient cohorts with chronic IMIDs.

Drawing on these concepts, here we present a novel systems genomics workflow that addresses the challenges of missing regulation and genetic epistasis, to predict the impact of non-coding SNPs on downstream cellular signalling networks as well as gene regulatory networks. We illustrate the utility of this approach by performing a patient-specific analysis in a well-powered cohort of 2636 individuals with Crohn’s disease (CD) and ulcerative colitis (UC), which represent two distinct but related chronic IMIDs that fall within the umbrella of IBD. CD and UC arise due to complex interactions between multiple genetic and environmental risk factors, resulting in the dysregulation of immune pathways in the gastrointestinal mucosa^21^. Using this workflow, we find that signals from disease-associated non-coding SNPs cumulatively propagate towards major biological pathways known to be pathognomonic to CD and UC at the signalling and gene regulatory layers. Importantly, SNP-propagated gene regulatory networks stratify CD and UC patients into distinct clusters, thereby capturing disease heterogeneity in IBD. We show that these genotype-driven patient clusters correspond to cell type-specific gene dysregulation and correlate with therapeutic response to biologic therapy. Thus, by modelling the impact of non-coding genetic variants on downstream cellular signalling pathways and gene regulatory networks using systems genomics, we bridge the gap between genotype and molecular phenotype in the prototypical complex genetic disorders of CD and UC. This approach has the potential to accelerate precision medicine efforts in IBD as well as other chronic IMIDs.

## Results

### Generation of patient-specific SNP-propagated signalling networks and gene regulatory networks

We used network propagation to unravel how disease-associated non-coding SNPs impact downstream cellular signalling and gene regulatory networks. We contextualised this analysis to the prototypical chronic IMIDs of UC and CD by utilising genotype data from a large cohort of genotyped CD (n=1,695) and UC patients (n=941) from the Leuven IBD Center as the input data for the workflow (University Hospitals Leuven, KU Leuven, Belgium, Table S1). The workflow, which is disease agnostic, is summarised in Figure 1. Briefly, we first annotated known CD and UC SNPs to non-coding regions of the genome (*i.e.* enhancers, promoters or miRNA-target sites) using our previously described iSNP method^17^. A gene and its translated protein were called “SNP-affected” if a non-coding IBD-associated SNP was predicted to modulate its expression by changes in the transcription factor binding site of the gene (in promoter or enhancer regions), or microRNA-target sites (miRNA-TS) that are in first intronic regions or untranslated regions (Table S1). The non-coding SNP-affected proteins were then assigned as “seed nodes” from which heat-based network propagation identified downstream nodes that could perturb or interact with in the global human intracellular signalling network ^22^. To identify the biological processes impacted in this “SNP-propagated signalling network”, we identified and annotated functional network units called modules. A network module is a part of the network which is more connected to its members than to other parts of the network, and often represent particular biological functions.

**Figure 1.**
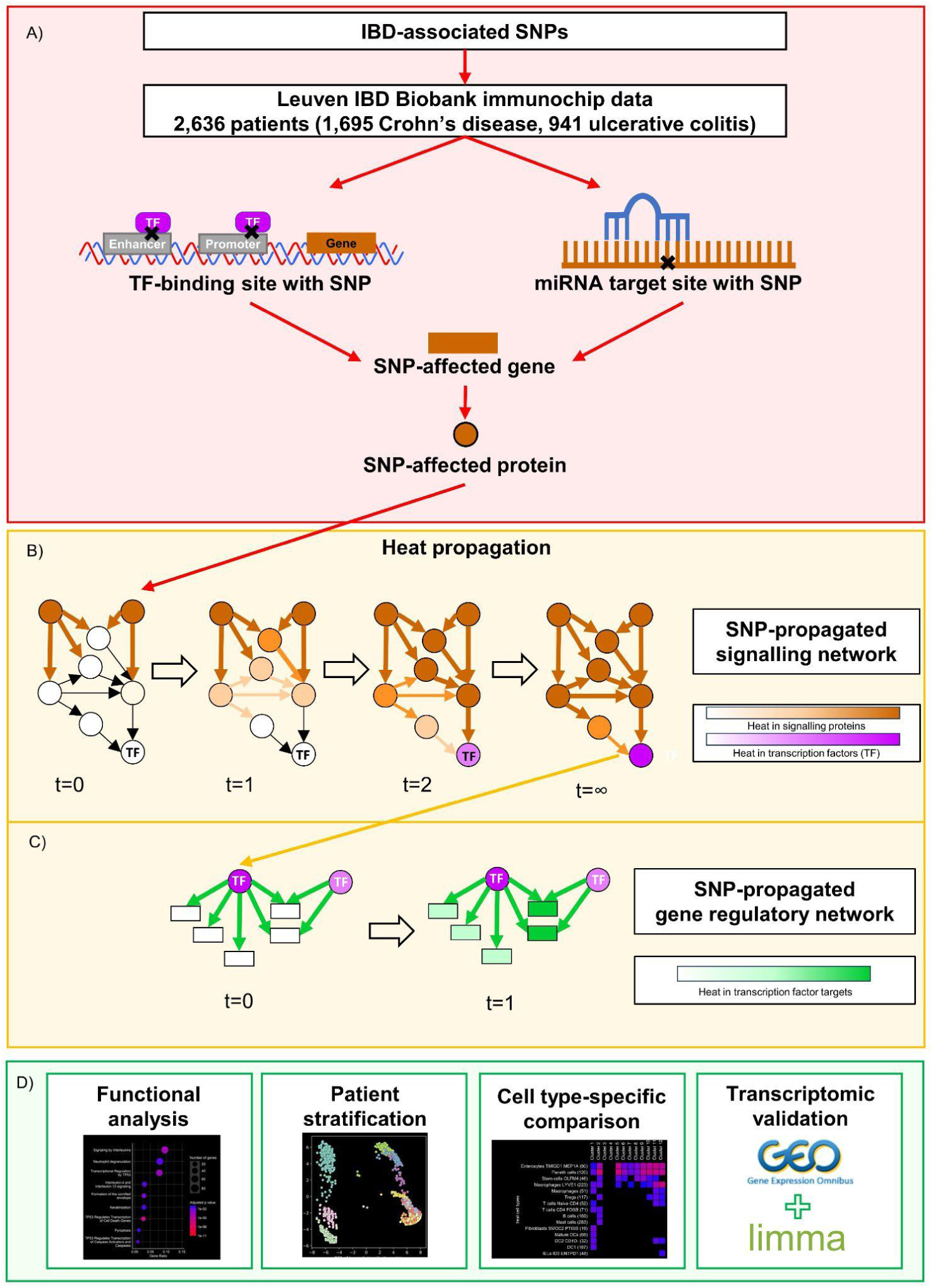
Workflow of the current systems genomics study. **A)** We first analysed genotype data from 2,636 IBD patients (1,695 Crohn’s disease, 941 ulcerative colitis) to evaluate the impact of non-coding IBD-associated SNPs on SNP-affected proteins based on the methodology of our previous work ^17^. **B)** Then, we utilised network heat propagation to determine the impact of non-coding SNPs beyond local first-neighbour SNP-affected proteins to downstream proteins to generate a SNP-propagated signalling network. This allowed us to better capture the impact of non-coding SNPs on the global intracellular signalling network. **C)** In this perturbed SNP-propagated signalling network, we then identified the downstream transcription factors, from which, using a further iteration of network heat propagation, we mapped the impact of non-coding SNPs on gene regulatory interactions. We defined this as the SNP-propagated gene regulatory network. **D)** Using the SNP-propagated signalling networks and gene regulatory networks, we performed various analyses including functional analysis, patient stratification, cell type-specific gene dysregulation comparison, and validation using bulk transcriptomics datasets. t: pseudotime, TF: transcription factor

We extended our analysis further by propagating the effect of non-coding SNPs through the downstream gene regulatory layer comprising transcription factor-target gene interactions ^23^. This gene regulatory network formed modules which were centred around major transcription factors and contained the target genes whose expression could be indirectly altered by disease-associated non-coding SNPs. Annotation of these gene regulatory modules allowed us to predict how CD and UC non-coding SNPs may perturb biological pathways and processes through their impact on the downstream gene regulatory layer.

In summary, these network propagation steps helped to enrich and contextualise non-coding genomic risk variants to intracellular processes that may contribute to pathomechanisms relevant in CD and UC. A summary of the features (non-coding SNPs, SNP-propagated genes, and SNP-propagated proteins) per propagation step for both CD and UC is provided in Figure S1.

### Non-coding SNPs in CD impact downstream signalling processes involving key immune pathways, DNA damage, and autophagy

We found 38 SNP-affected proteins in our cohort of 1,695 CD patients (Table S2). We used these proteins as seeds for the heat diffusion algorithm to reconstruct a SNP-propagated signalling network comprising 518 proteins (nodes) and 1,462 signalling interactions (edges). The network consisted of five major modules (Figure 2A, Table S3). The key biological pathways enriched in these modules included those relating to immune pathways (mast cell chemotaxis, tyrosine-kinase signalling including JAK-STAT regulation, IL-2 regulation, MAPK and NFkB regulation), DNA damage (cell cycle arrest, DNA damage checkpoints, apoptosis), and autophagy. These findings reveal how the genetic background of patients may be poised to impact major signalling processes that are known to be involved in CD pathogenesis.

**Figure 2.**
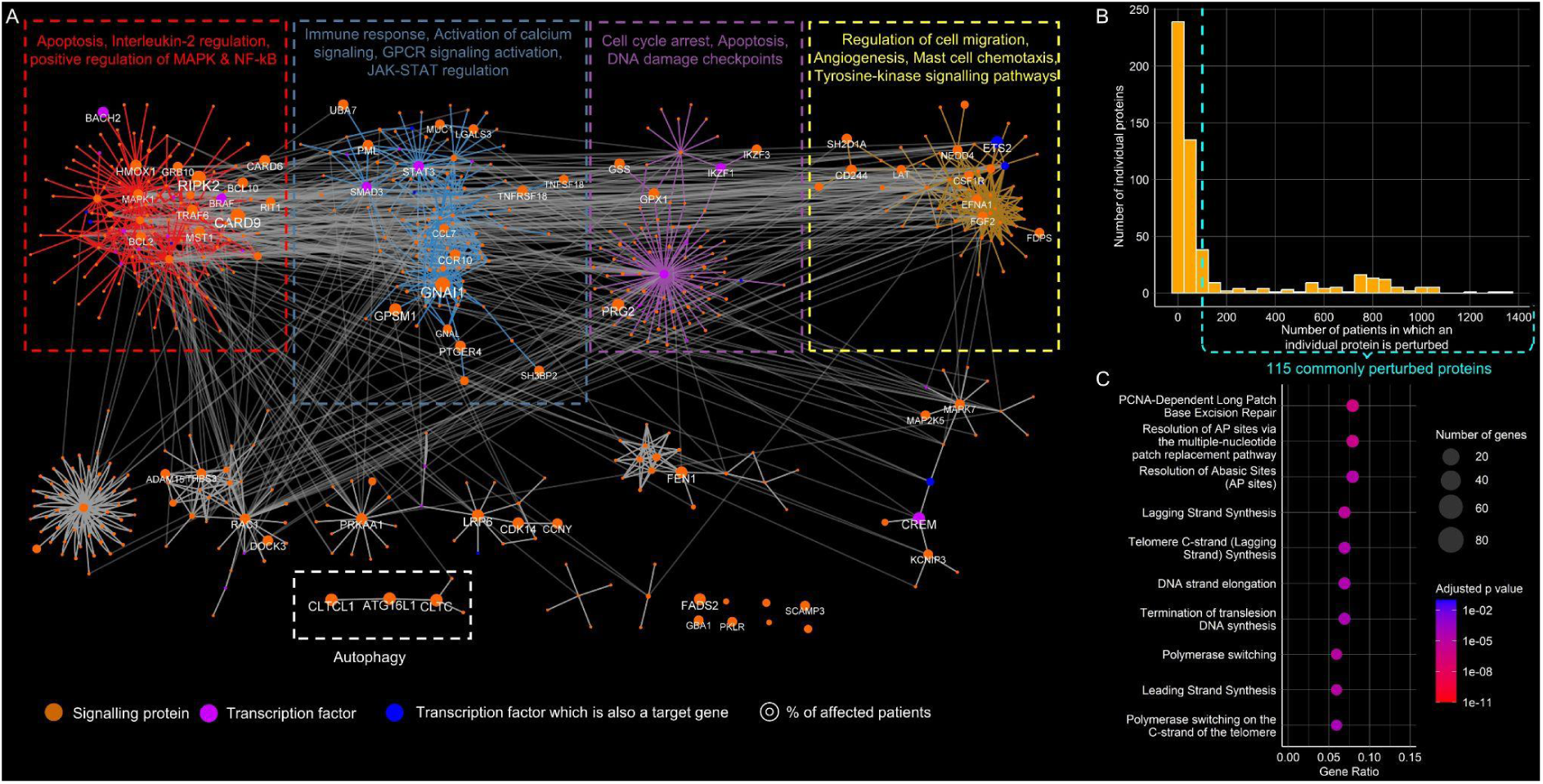
Crohn’s disease-associated non-coding SNPs perturb the cell cycle, autophagy, apoptosis and immune responses. **A)** SNP-propagated signalling network in Crohn’s disease (CD) patients grouped by Girvan–Newman modularisation. The largest four modules were functionally annotated using Reactome enrichment. Node size corresponds to the number of patients having the SNP-propagated protein. **B)** Frequency distribution of SNP-propagated proteins in the CD patient cohort. **C)** The enriched Reactome pathways of the most commonly perturbed SNP-propagated proteins in the CD patient cohort. Commonly perturbed proteins are those that are affected in more than 100 patients in the cohort.

Non-coding SNPs were predicted to impact 115 proteins in more than 100 CD patients (Figure 2B). These proteins were enriched in DNA synthesis and DNA repair pathways in Reactome ^24^ due to the commonly perturbed polymerase delta subunits (POLD1,2,3,4) and their cofactors replication factor C (RFC2,3), known to be involved in DNA damage repair and replication ^25^. This suggests that non-coding SNPs may influence the maintenance of genomic integrity in a significant subset of CD patients (Figure 2C, Figure S2A). Additional putative perturbed signalling pathways included cell-cell communication related processes, such as interleukin signalling and cell adhesion (Table S4), which have been previously described in CD pathogenesis ^26,27^. The commonly perturbed proteins did not significantly overlap with differentially expressed genes when comparing CD patients and non-IBD controls from previous studies (Benjamini Hochberg adjusted p> 0.05, Table S5).

### Patient-specific SNP-propagated gene regulatory networks stratify CD patients into 12 clusters

We hypothesised that propagating the signal further through a downstream transcription factor-target gene regulatory network may better capture the differences in gene expression between CD patients and non-IBD controls. The CD SNP-propagated gene regulatory network comprised 2,093 genes in total of which 26 were transcription factors, 2,056 were target genes, and 11 were both transcription factors and target genes. These genes formed 3,728 transcription factor-target gene (TF-TG) interactions. The TF-TG network formed seven gene regulatory modules (Figure 3A). The regulatory modules corresponded to various CD-associated functions: i) MAPK signalling (regulated mainly by TFs such as ETS1, EST2, and FLT1); ii) B cell receptor and T cell receptor signalling and Rho GTPase cell cycle (regulated by IKZF1); iii) p53 regulated genes, iv) hemidesmosome assembly and interleukin signalling (regulated by various TFs relevant in other modules as well); v) neutrophil degranulation, regulation of complement cascade, and IL4 and IL13 signalling (regulated by BRAF and STAT3); vi) integrin cell surface interactions and TGFB signalling (regulated by SMAD3 and ZFP36); and vii) signalling by IL7 and other interleukins (regulated by multiple transcription factors including FOXP3) (Table S3). These findings suggest that disease-associated non-coding SNPs may disrupt key pathogenic pathways in CD through their cumulative influence on the downstream signalling and gene regulatory layers.

**Figure 3.**
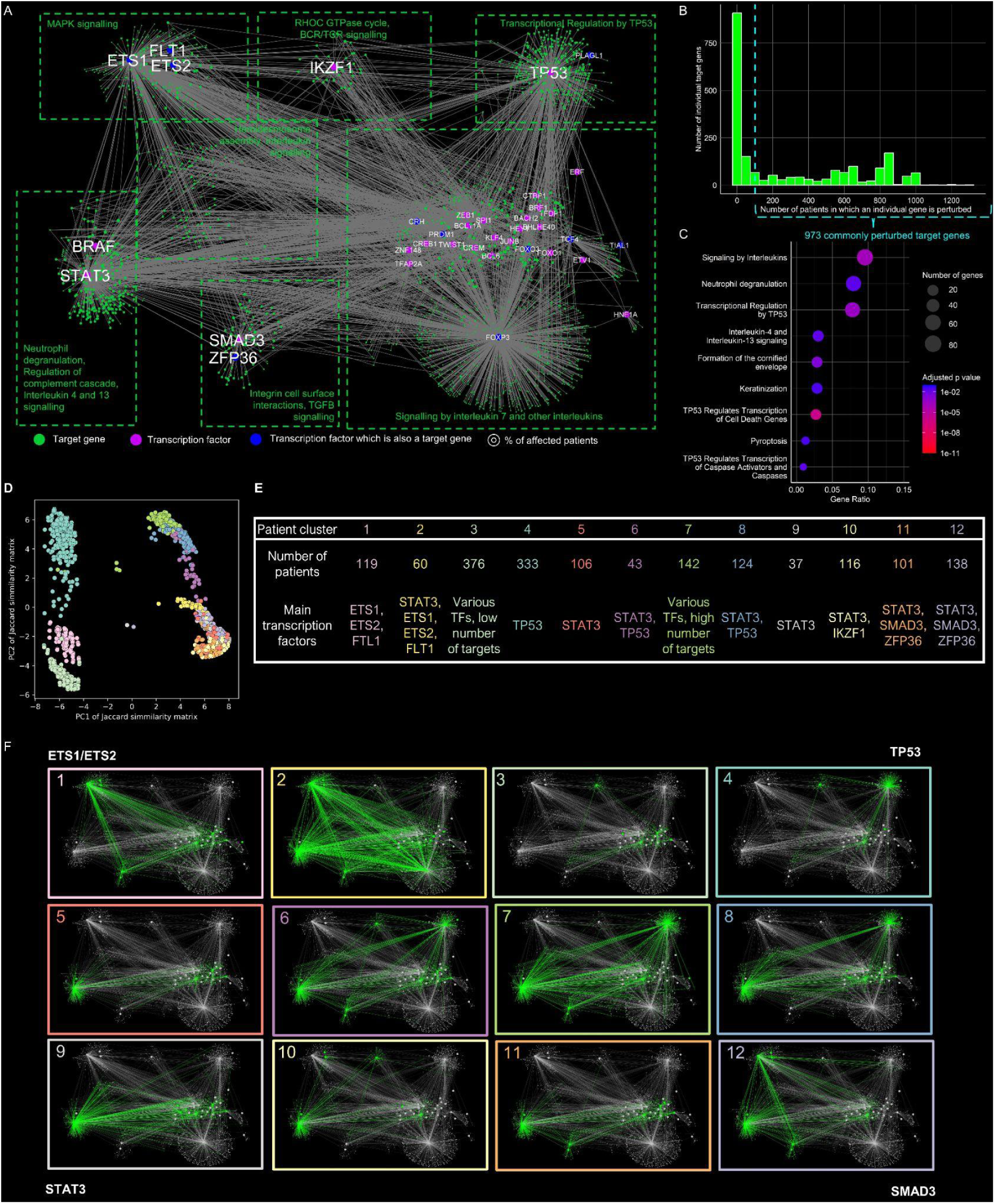
SNP-propagated gene regulatory network divides patients into 12 clusters in Crohn’s disease. **A)** The SNP-propagated gene regulatory network in Crohn’s disease (CD). Pink nodes represent transcription factors, while green nodes are target genes of the transcription factors, and blue nodes are both. Size of target gene nodes corresponds to the number of patients having the SNP-propagated target genes. The network represents patient-specific gene regulatory modules. **B)** Frequency distribution of SNP-propagated transcription factor-target genes in the CD patient cohort. **C)** The enriched Reactome pathways of the commonly perturbed SNP-propagated transcription factor-target genes in the CD patient cohort. **D)** Principal component analysis of the Jacquard similarity matrix between the patient-specific gene regulatory networks. Each node represents a patient and the colours are according to k-medoid clustering where k is 12. **E)** Patient-specific clusters and their cluster-determining SNP-propagated transcription factors. **F)** Representative examples of CD patient-specific regulatory networks.

The majority of the target genes in the CD SNP-propagated gene regulatory network were present in small numbers of patients (Figure 3B). However, 973 target genes were predicted to be perturbed in more than 100 patients. These commonly perturbed genes significantly overlapped with CD-specific differentially expressed genes, both in the ileum and rectum (p< 0.05 Benjamini-Hochberg corrected p-value, Table S5), and were functionally enriched for interleukin signalling, P53-related processes, and neutrophil degranulation (Figure 3C, Supplementary Figure 2C, Table S4). Interleukin signalling and DNA damage repair processes were also identified in the SNP-propagated regulatory network as enriched pathways similarly to the upstream SNP-propagated cell signalling network. This overlap suggests that SNP-propagated pathogenic feedback loops may connect the cell signalling and gene regulatory layers of these pathways in CD. The enrichment of neutrophil degranulation, which is a characteristic feature of active IBD ^28^ and the basis of faecal calprotectin testing as a non-invasive biomarker ^29^, indicates a potential genetic mechanism contributing to this phenomenon.

We next evaluated whether the SNP-propagated networks could dissect patient heterogeneity in our cohort through clustering analysis. The clustering of SNP-propagated gene regulatory networks yielded a higher average silhouette score compared to the SNP-propagated signalling network and the SNP-affected proteins, suggesting that patient heterogeneity is most effectively captured at the gene regulatory layer (Supplementary Figure 3). Using k-medoid clustering, the patient-specific SNP-propagated gene regulatory networks stratified the CD cohort into 12 distinct patient-specific clusters (Figure 3D, Figure S3). Each patient cluster was characterised by a corresponding set of gene regulatory modules driven by the major transcription factors ETS1/ETS2 (patient clusters 1, 2, 7), TP53 (patient clusters 4, 6, 7, 8), STAT3 (patient clusters 2, 5, 6, 7, 8, 11,12), and SMAD3 (patient clusters 7,11,12) (Figure 3E and 3F). This analysis indicates that there is a spectrum of different gene regulatory interactions that may be impacted by non-coding SNPs in different clusters of CD patients. Determining the patient-specific differences and similarities between these gene regulatory networks could provide a better understanding of how patient-specific combinations of non-coding SNPs contribute to heterogenous pathomechanisms in CD. Similar insights have already been observed in cancer, in which gene regulatory networks stratified patients with various types of cancer into distinct subgroups with prognostic implications ^30^.

### Patient-specific clusters of SNP-propagated gene regulatory modules correspond to cell type-specific gene dysregulation patterns in CD patients

We posited that the identified CD patient clusters of SNP-propagated gene regulatory modules may have cell type-specific effects. To validate this prediction, we used two large publicly available single-cell transcriptomics dataset (Kong *et al* 2023^31,32^ and Krzak *et al* 2024^31,32^) ^31,32^ of CD patients (n=46 and n=26 respectively) and healthy controls (n=25 and n=25 respectively) to determine if the target genes in each of the 12 patient-specific clusters of SNP-propagated regulatory networks corresponded to CD single-cell differentially expressed genes (DEGs). In both datasets, the patient-specific clusters mapped to differential gene expression in a cell type-specific manner that was also location dependent (Figure 4, Figure S4, Table S6). In the Kong *et al* cohort, the majority of CD patient clusters corresponded to the DEGs between non-inflamed CD and healthy samples present in enterocytes, Paneth cells, and stem cells located in the ileum. In particular, clusters 1 (regulated by ETS1, ETS2, and FTL1) and 2 (STAT3, ETS1, ETS2 and FTL1) corresponded exclusively to DEGs originating from multiple ileal epithelial and immune cell types.. Only cluster 3 patients mapped to DEGs from colonic cell types alone, whilst certain clusters such as clusters 11 and 12 (both regulated mainly by STAT3, SMAD3, and ZFP36) mapped to cell types present in both the ileum and colon. Of note, cluster 9 (which was regulated by STAT3) corresponded to DEGs in goblet cells present in colonic tissues, suggesting that these patients may have altered goblet cell function. Due to the paucity of the available clinical meta-data, it is unclear whether these location-specific differences reflect differences in CD phenotype (i.e. ileal vs colonic vs ileocolonic disease). However, findings from one of the largest genotype-to-phenotype studies in IBD to date, indicate that ileal and colonic CD may represent genetically distinct clinical entities ^33^. In the independent Krzak *et al* cohort (Figure S5A), the majority of clusters (9/12) corresponded to DEGs in enterocytes, goblet cells, naive B cells and memory B cells. Clusters 2 and 9 (both regulated by STAT3) appeared to be the most pervasive clusters affecting most cell types. Only SNP-propagated gene regulatory networks in Cluster 4 (regulated by p53) failed to correspond to ileal or colonic cell type-specific gene expression signatures in both these cohorts of CD patients.

**Figure 4.**
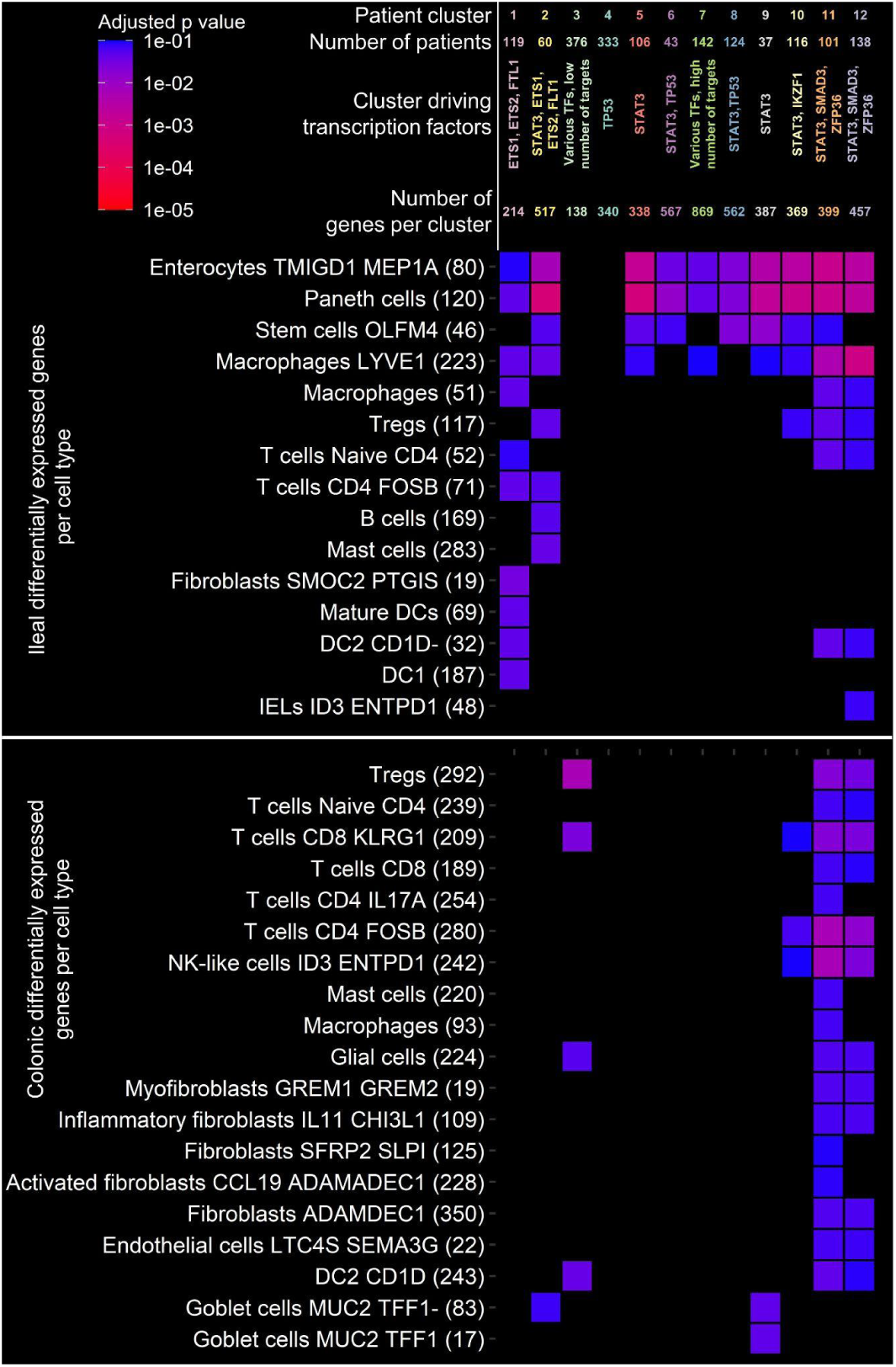
Patient-specific clusters correspond to cell type-specific differentially expressed genes in Crohn’s disease. The x axis corresponds to the various patient-specific clusters based on the SNP-propagated gene regulatory networks. The y axis denotes intestinal cell types with differentially expressed genes (DEGs) from Crohn’s disease (CD) patients. DEGs were defined comparing the expression of healthy vs non-inflamed CD biopsies from the Kong *et al.* study (|FC| >1, Benjamini-Hochberg corrected Wilcoxon rank sum test p<0.1 ^31^). In brackets the number of differentially expressed genes between healthy and non-inflamed CD cells in a given cell type is shown. If a square has a colour then the cluster-specific genes are overrepresented in the cell type-specific differentially expressed gene set (Benjamini-Hochberg adjusted hypergeometric test p<0.1). If a square is black, that means a cluster is not overrepresented in the cell type-specific CD signature. FC: Fold change

We also validated the transcription factors predicted to be perturbed in the CD patient clusters of SNP-propagated gene regulatory modules by inferring transcription factor activity from the gene expression data of CD patients from the Kong *et al* and Krzak *et al* studies (Figure S6 S5B). Strikingly we found that out of the 6 major cluster-representative transcription factors (ETS1, ETS2, STAT3, IZKF1, SMAD3, and P53) which had more than 5 target genes, all six were predicted to have perturbed activities in the Kong *et al* dataset. In the Krzak *et al* dataset, 5 out of the 6 transcription factors were predicted to have altered activities. Again, there were notable differences in transcription factor activities between ileal and colonic tissues in both datasets. STAT3, which is one of the main tightly regulated downstream transcription factors in cytokine signalling ^34^, was upregulated in almost all cell types in the Krzek *et al* dataset and in ileal immune cell types in the Kong *et al* dataset. Meanwhile SMAD3 had a strong stromal signal and was upregulated in myofibroblasts in both datasets. In summary, we found that signals from CD-associated non-coding SNPs propagate towards downstream gene regulatory networks through certain transcription factors in a cell type- and location-specific manner.

### In UC, non-coding SNPs impact signalling processes involving adaptive immune pathways, cytokine signalling, fatty acid metabolism, and chemotaxis

We next investigated whether non-coding SNPs impart similar or distinct effects in UC compared to CD. We identified 38 non-coding SNP-affected proteins in our cohort of 941 UC patients (Table S2). Similar to the CD analysis, we used these proteins as seeds for the heat diffusion algorithm and reconstructed a large SNP-propagated signalling network containing 348 proteins and 1,077 protein-protein interactions. This UC SNP-propagated signalling network comprised four major functional modules (Figure 5A). The Reactome pathway analysis revealed that the key cellular processes enriched in these modules were related to: i) MHC Class II antigen presentation, interferon gamma signalling, and T cell receptor activation; ii) NFkB, toll-like receptor, IL1 and IL17 signalling; iii) fatty acid metabolism and IL10 signalling; and iv) defence response and chemotaxis (Table S3). Most of these pathways are known to be involved in UC pathogenesis ^35^, although the role of fatty acid metabolism is an area of ongoing investigation ^36^.

**Figure 5.**
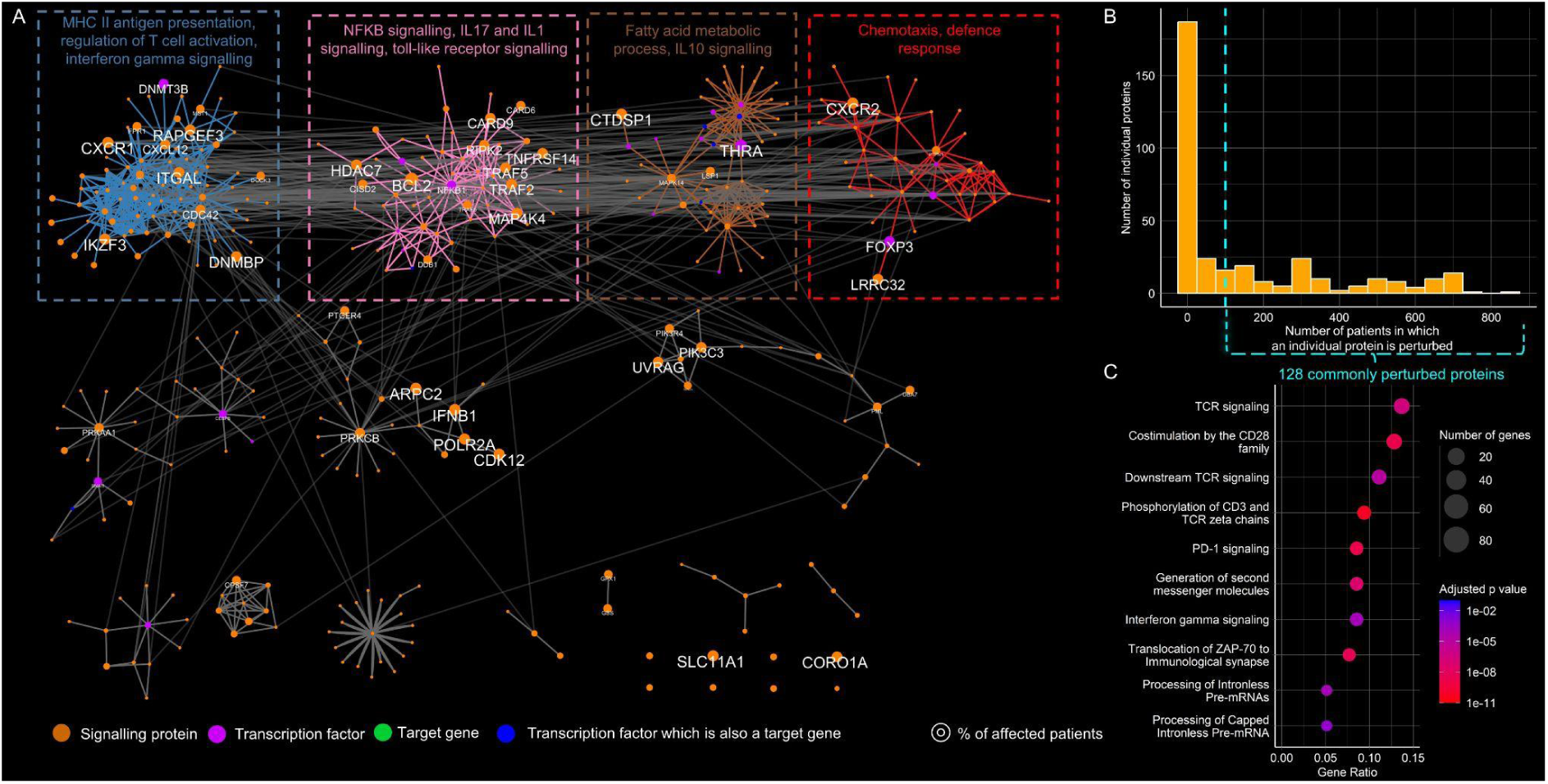
Ulcerative colitis-associated non-coding SNPs affect T-cell activation, NFKB signalling, cytokine signalling, and chemotaxis. **A)** SNP-propagated signalling network in ulcerative colitis (UC) patients grouped by Girvan–Newman modularisation. The largest four modules functionally annotated using Reactome enrichment. Node size corresponds to the number of patients harbouring the SNP-propagated protein. **B)** Frequency distribution of SNP-propagated proteins in the UC patient cohort. **C)** The enriched Reactome pathways of the most commonly affected SNP-propagated proteins in the UC patient cohort.

The vast majority of SNP-propagated signalling proteins were relevant only in a small number of patients. However, 128 proteins were predicted to be perturbed in at least 100 UC patients in our cohort (Figure 5B). This distribution was similar to our observations in CD. The commonly perturbed proteins in UC were functionally enriched for pathways related to T cell receptor activation/signalling and the interferon gamma pathway (Figure 5C, Figure S2C, Table S4). As these are established pathogenic pathways in UC ^35^, the systems genomics approach presented here provides novel insights into the molecular mechanisms linking non-coding UC-associated risk variants with known pathogenic drivers of the disease. These SNP-propagated proteins were significantly enriched in the transcriptomics signature for inflamed UC samples but not non-inflamed samples, suggesting that the SNP-propagated signalling network is more relevant in UC pathogenesis in the context of active rather than quiescent disease (Table S5).

### In UC, patient-specific combinations of SNP-propagated gene regulatory modules are driven by four transcription factors that stratify patients into seven clusters

We next evaluated whether distinct pathogenic pathways could be perturbed by non-coding SNPs at the downstream gene regulatory layer compared to the signalling layer in UC, similar to what we had observed with CD. The UC SNP-propagated gene regulatory network consisted of 3,372 genes and 4,979 TF-TG interactions. The network was formed by 20 transcription factors, 3,346 target genes, and 6 genes which functioned as both. Six gene regulatory modules were identified in this regulatory network (Figure 6A). These modules corresponded to various biological functions: i) retinoid acid and bile acid metabolism (regulated mainly by SNAI1, CEPBP, RXRA and other TFs); ii) xenobiotics (regulated by multiple TFs such as ETS1, ETS2, AHR, FOS); iii) NFkB1, interleukin, and toll-like receptor signalling (regulated by NFkB); iv) cell cycle regulation (regulated by E2F1, E2F2, E2F3, and NOTCH4); and v) NOTCH1 and NOTCH2 signalling (regulated mainly by EGR1). A sixth module was regulated by FOXP3, but its target genes were not significantly enriched with any pathways (Table S3). Thus, as seen in CD, the UC SNP-propagated regulatory network analysis revealed that non-coding SNPs perturb different but highly relevant pathogenic pathways at the gene regulatory layer compared to those at the signalling layer in UC.

**Figure 6.**
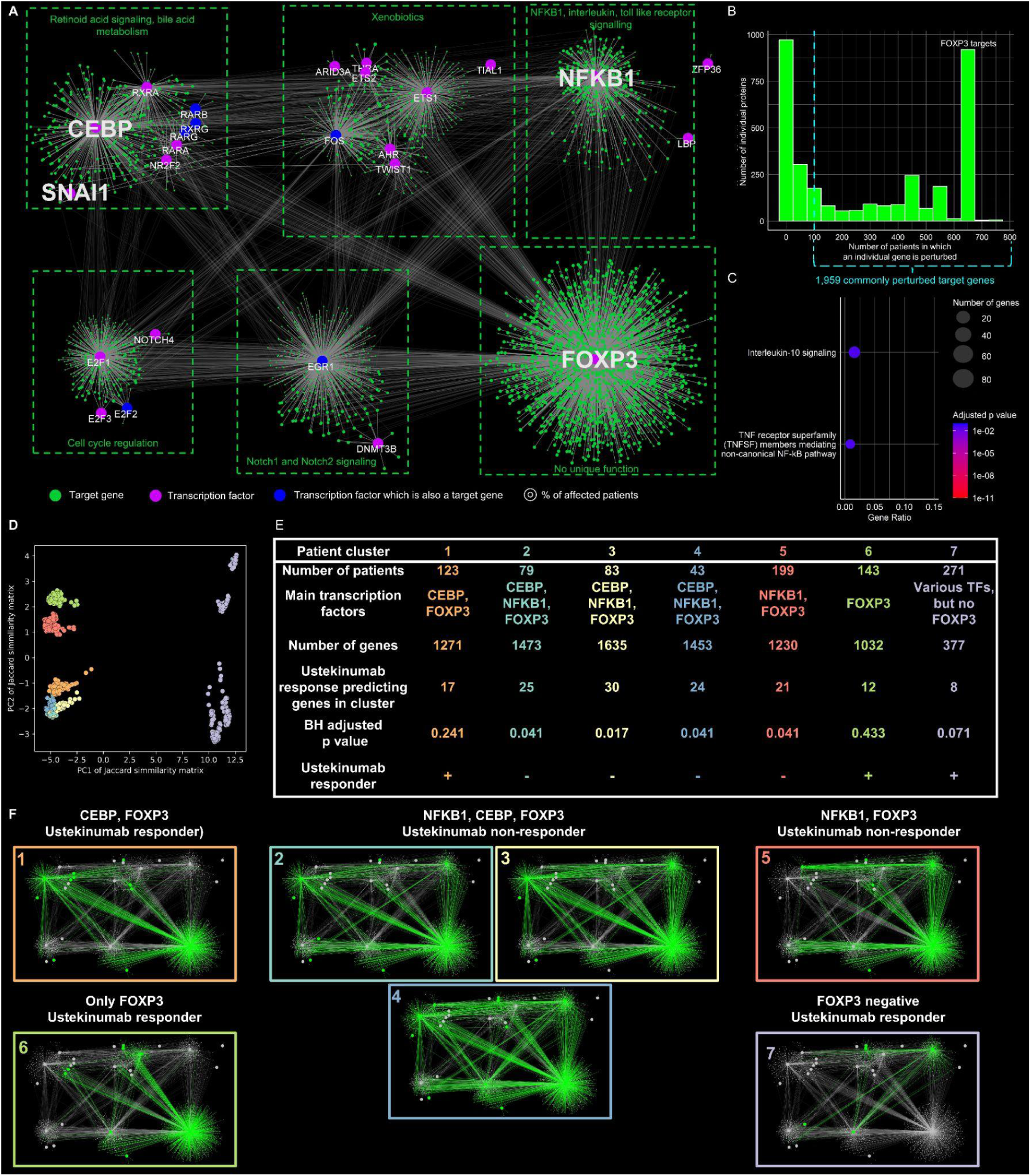
SNP-propagated gene regulatory network divides ulcerative colitis patients into 7 clusters based on three main transcriptional regulators: CEBP, NFKB1 and FOXP3. **A)** The SNP-propagated gene regulatory network in ulcerative colitis (UC) patients. Pink nodes represent transcription factors, while green nodes are transcription factor target genes, and blue nodes are both. Size of target gene nodes represents the number of patients having the SNP-propagated target genes. **B)** Frequency distribution of SNP-propagated transcription factor-target genes in the UC patient cohort. **C)** Significantly enriched Reactome processes in the most commonly perturbed SNP-propagated target genes in the UC patient cohort. **D)** Principal component analysis of the Jaccard similarity matrix between the patient-specific gene regulatory networks. **E)** Patient-specific clusters and their cluster-determining transcription factors. The enrichment of DEGs comparing ustekinumab response vs non-response is also shown here. **F)** Representative examples of UC patient-specific SNP propagated gene regulatory networks.

Unlike the UC SNP-propagated signalling network, the frequency distribution of target genes in the SNP-propagated regulatory network had a bimodal distribution (Figure 6B) - one peak with fewer than 50 patients, and the other peak involving 669 patients. The latter peak comprised the target genes of FOXP3. We next identified target genes that were perturbed in at least 100 UC patients in our cohort. Similar to the UC SNP-propagated signalling network, these 1,959 commonly perturbed target genes were enriched for the transcriptomics signature of inflamed UC intestinal tissues but not non-inflamed samples (Table S5). Functional annotation of these 1,959 genes revealed that they were involved in IL10 and non-canonical NFκB signalling mediated by TNF receptor superfamily members (Figure 6C, Table S4). This indicates that in a significant subset of UC patients, there is a dysregulation of critical pro- and anti-inflammatory pathways as a consequence of their underlying genetic background, which is likely to be relevant in the context of active inflammation. These findings further underscore the significance of understanding how non-coding disease-associated SNPs disrupt the downstream gene regulatory layer.

We next evaluated whether we could stratify the UC patients in our cohort using the SNP-propagated signalling or gene regulatory networks. Similar to our observations in CD, clustering of the SNP-propagated gene regulatory network performed best for UC patient stratification (Figure S3), resulting in 7 distinct patient clusters (Figure 6D). This stratification was based on the combination of involved gene regulatory modules specifically driven by FOXP3, NFKB1, CEBP, and SNAI1 (Figure 6E and 6F). Unlike CD, however, the UC SNP-propagated patient clusters did not correspond with UC cell type-specific signatures obtained from the Smillie *et al* single-cell dataset ^37^. This could be due to the low power of the Smillie *et al* dataset (n=17 UC patients) compared to the Kong *et al* dataset (n=46 CD patients).

### Patient-specific clusters of SNP-propagated gene regulatory modules correspond to therapeutic response to ustekinumab therapy in UC patients

To understand whether the stratified UC patient clusters correspond to clinically relevant phenotypes, we investigated whether they may influence therapeutic response. To evaluate this, we utilised data from a randomised placebo-controlled study called UNIFI, which investigated the efficacy and safety of ustekinumab (anti-IL-12/IL23 p40 antibody) for the treatment of moderate-to-severe UC ^38^. In comparing responders and non-responders only patient clusters 2, 3, 4 and 5 were enriched in differentially expressed genes (Benjamini-Hochberg corrected hypergeometric test p<0.05, Figure 6E and 6F). These clusters were characterised by both NFKB1- and FOXP3-driven gene regulatory modules, while the remaining patient clusters were driven by either one or none of these transcription factors. Due to this, the patients in clusters 2, 3, 4 and 5 may have a lower chance of responding to ustekinumab therapy, because their genetic background perturbs the genes which differentiate between treatment response and non-response. Whilst the precise mechanisms need further experimental evaluation, these findings suggest a hitherto unrecognised genetic basis for ustekinumab therapeutic response in UC patients, which could be leveraged for future precision medicine strategies.

### CD and UC non-coding SNPs converge on the same signalling pathways, but they perturb distinct gene regulatory modules

After separately analysing non-coding SNPs from CD and UC patients, we compared the two forms of IBD with each other. We created a combined IBD SNP-propagated signalling network and a combined IBD SNP-propagated gene regulatory network. The combined IBD SNP-propagated signalling network revealed three major modules of perturbed signalling proteins, of which two corresponded to T cell activity/GPCR activity and inflammation/apoptosis/cell cycle/NFkB1 regulation (Figure 7A). We identified that different signalling interactions link CD and UC SNPs to key signalling proteins such as ETS2 and CARD9. The common proteins in the combined IBD SNP-propagated signalling network were enriched for interleukin signalling and T cell receptor signalling (Figure 7B). The combined SNP-propagated signalling network was also composed of small, disease-specific network modules such as the autophagy module in CD or the module involving CEBPB in UC, suggesting that these processes are likely to have disease-specific genetic associations from multiple SNPs.

**Figure 7.**
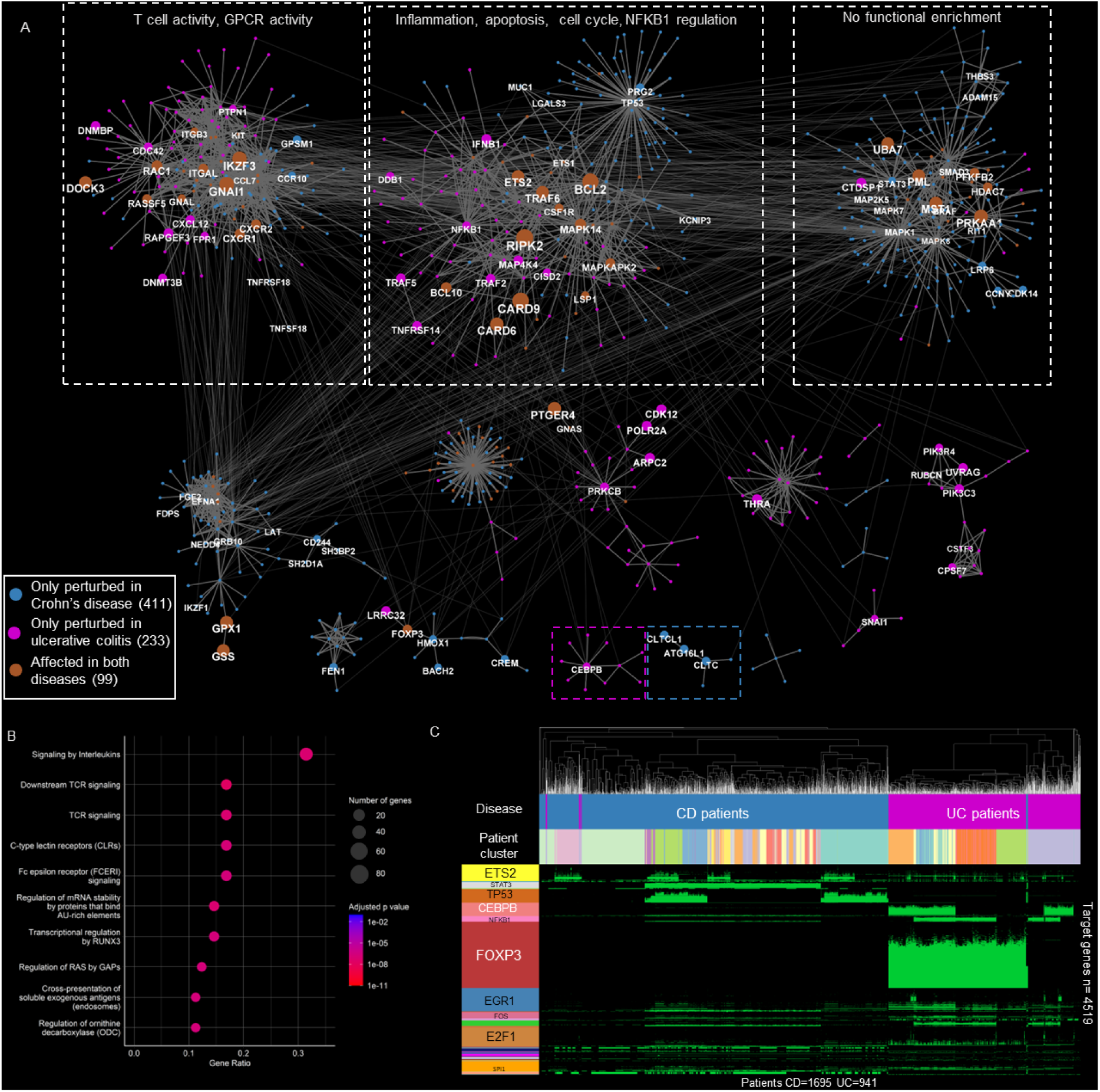
Crohn’s disease and ulcerative colitis patients share several pathways in their SNP-propagated signalling networks, whilst their SNP-propagated gene regulatory networks reveal distinct transcription factor-target gene interactions. **A)** Combined SNP-propagated signalling network of Crohn’s disease (CD) and ulcerative colitis (UC) patients. Node size corresponds to the number of CD or UC patients having the SNP-propagated protein. Two disease-specific modules are highlighted: CEBPB regulation in UC (pink) and autophagy in CD (blue). **B)** Reactome pathway enrichment of the SNP-propagated proteins perturbed in both diseases. **C)** Heatmap of SNP-propagated transcription factors and their target genes in CD and UC patients. The columns represent patients clustered according to the transcription factor-target genes present in the SNP-propagated gene regulatory network in CD (blue) and UC (purple). Rows represent SNP-propagated transcription factors. Within the heatmap, green colour indicates transcription factor-target gene interactions present within the SNP-propagated gene regulatory network, whilst black indicates interactions that are not present.

Unlike the SNP-propagated signalling network, the combined IBD SNP-propagated gene regulatory network revealed clear differences between CD and UC. This was because signals propagated by non-coding SNPs perturbed downstream TFs which have little overlap between CD and UC, leading to distinct downstream gene regulatory interactions (Figure 7C, Figure S7). STAT3 and TP53 were identified as TFs specifically perturbed in CD, whilst CEBPB, FOXP3, and NFκB1 were TFs specifically perturbed in UC. The only overlap we observed was in ETS2-regulated target genes which were perturbed in both CD and UC SNP-propagated gene regulatory networks.

## Discussion

In this study, we present a novel systems genomics workflow that addresses the challenges of missing regulation and genetic epistasis to disentangle the putative biological mechanisms linking non-coding genetic variants to pathogenic pathways in complex diseases. Previous studies have focused on integrating GWAS signals with chromatin accessibility, expression quantitative trait loci (eQTLs), and gene expression data ^39^, or mapping SNP-affected genes to PPI networks ^16,40,41^ to identify disease-relevant genes or pathways. However, these approaches do not consider the potential downstream impact of genetic risk loci on both the signalling and gene regulatory layers. This would be critical to understanding how non-coding genetic risk variants ultimately impact pathogenic pathways and influence phenotype in complex diseases such as chronic IMIDs. Hence, we leveraged network propagation to model the holistic impact of disease-associated non-coding SNPs, extending beyond local interacting neighbours to encompass the global topology of cellular signalling and gene regulatory networks.

Using this methodology, we performed the largest patient-specific systems genomics analysis in chronic IMIDs thus far, by utilising genotype data from over 2500 individuals with the two major forms of IBD: CD and UC ^21,42^. Despite the success of GWAS in identifying over 240 susceptibility variants in IBD to date^42^, genotype-phenotype associations have been disappointingly sparse both in terms of genotype-driven pathomechanisms and genotype-associated clinical outcomes ^33^. Similar observations have been made in multiple chronic IMIDs such as rheumatoid arthritis, multiple sclerosis, and type 1 diabetes ^5^. As a result, the initial optimism surrounding the use of genomics for identifying patient-specific pathomechanisms and advancing precision medicine efforts in chronic IMIDs has been dampened, prompting many in the field to shift their focus to other -omic layers ^43^. In this work, however, we were able to unravel the heterogenous signalling and gene regulatory mechanisms connecting individual patient genotypes with phenotypes in the prototypical chronic IMIDs of UC and CD. In doing so, we demonstrate that patient genotype can offer crucial biological and translational insights which have potential to inform future personalised medicine approaches. This is urgently required given the significant therapeutic ceiling effect currently observed across multiple chronic IMIDs ^44–46^.

The presented analysis revealed that UC- and CD-associated non-coding genetic variants propagate towards and perturb key pathogenic pathways known to be implicated in IBD pathogenesis at both the downstream signalling and gene regulatory layers. For instance, at the signalling layer, CD-associated non-coding SNPs were predicted to impact proteins involved in apoptosis ^47,48^ and autophagy ^49,50^, which are both known drivers of CD pathogenesis (Figure 2). Interestingly, while autophagy dysfunction in CD is traditionally attributed to the *ATG16L1* coding risk variant ^51^, our findings suggest that autophagy signalling may be disrupted in patients possessing two non-coding SNPs (rs3792112 and rs3828309) which propagate towards ATG16L1 and two other autophagy-related proteins (CLTC1 and CLTC). In UC, we were also able to predict how established UC-associated signalling processes such as NFκB signalling ^52^, cytokine signalling involving IFNɣ ^53^, IL1 ^54,55^ and IL17 ^56^, and MHC-II based antigen presentation ^57^, are impacted by UC-associated non-coding SNPs (Figure 5).

At the downstream gene regulatory layer, non-coding SNPs were predicted to impact several biological pathways implicated in IBD pathogenesis that were driven by key transcriptional regulators. In CD, these included neutrophil degranulation ^58^ (regulated by STAT3 and BRAF), integrin cell surface interactions ^59^ (regulated by SMAD3 and ZFP36), IL7 signalling^60^ (regulated by multiple transcription factors), and MAPK signalling ^61^ (regulated by ETS1, ETS2, FLT1) (Figure 3). In UC, retinoic acid signalling and bile acid metabolism ^62,63^ (regulated by CEBP, SNAI1, and other transcription factors such as NR2F2, RXRA), Notch signalling ^64,65^ (regulated by EGR1), and interleukin and toll-like receptor signalling (regulated by NFᴋB1), emerged as the key pathways likely to be impacted by non-coding SNPs at the gene regulatory layer (Figure 6). These findings illustrate how small regulatory changes influenced by non-coding SNPs contribute to IBD pathogenesis through mostly distinct pathways at the gene regulatory layer in UC and CD.

Combined analysis and visualisation of both UC and CD SNP-propagated signalling networks revealed that distinct non-coding SNPs associated with UC and CD can affect the same intracellular processes such as T cell activity, apoptosis, and NFkB1 regulation, but through different disease-specific signalling interactions (Figure 7A). Thus, at the signalling layer, non-coding SNPs ultimately converge on key pathways that are shared between the two forms of IBD but through different pathomechanistic routes. In contrast, little overlap was observed between the SNP-propagated gene regulatory networks of CD and UC patients (Figure 7C; Supplementary Notes), emphasising the distinct gene regulatory mechanisms underpinning CD and UC.

Consistent with these observations, we found that SNP-propagated gene regulatory networks stratified our patient cohort more effectively than either SNP-propagated signalling networks or neighbouring SNP-affected genes, emphasising the importance of harnessing the gene regulatory layer to dissect heterogeneity in IBD (Figure S3). The SNP-propagated gene regulatory networks stratified CD and UC patients into distinct patient clusters, revealing heterogenetic gene regulatory pathomechanisms across the various patient subgroups. In CD, we contextualised these clusters to specific cell types and spatial location along the gastrointestinal tract using single-cell transcriptomics data (Figure 4 and Figure S4,S5,S6). For instance, two SNP-propagated patient clusters regulated by ETS1 and ETS2 were enriched in certain immune (e.g. macrophages, CD4 T cells, and dendritic cells) and epithelial cell types (e.g. enterocytes and Paneth cells) predominantly in the ileum. This complements a recent finding that a pleiotropic non-coding risk variant associated with CD and other immune mediated disorders perturbs ETS2, which functions as a master regulator of pro-inflammatory responses in human macrophages ^66^ Although this variant was absent in our patient cohort, our findings suggest that other non-coding SNPs have potential to indirectly affect ETS2 function in a subset of CD patients, not only in macrophages but also in additional disease-relevant immune and epithelial cell types. Furthermore, in UC, we demonstrated that SNP-propagated gene regulatory networks corresponded to therapeutic response to ustekinumab (anti-IL-12/23 p40 monoclonal antibody), which is a widely used biologic in current clinical practice. Specifically, we observed that only UC patient clusters that had both NFkB1- and FOXP3-perturbed modules in their SNP-propagated gene regulatory networks were enriched for differentially expressed genes in ustekinumab responders and non-responders from the UNIFI trial ^38^ (see Supplementary Notes for further discussion) (Figure 6E and 6F). This reveals a previously unrecognised genetic basis for ustekinumab therapeutic response in UC patients, which could be harnessed for future precision medicine strategies.

Our findings open up several lines of future experimental enquiry to mechanistically interrogate the effects of non-coding SNPs in CD and UC on cell signalling and gene regulatory pathways in disease-relevant cell types. We believe this will be critical for achieving the goal of precision medicine in IBD and other IMIDs (please also refer to Supplementary Notes). Recent preliminary reports demonstrate that the field is already moving towards this direction by investigating the functional role of SNPs in disease-relevant cell types. This includes the mapping of the pleiotropic intergenic risk variant located on chr21q22 to *ETS2* dysfunction in inflammatory macrophages ^66^ and the identification of *CLN3* as a causal candidate gene for CD susceptibility in type 3 innate lymphoid cells ^67^. The workflow developed for this study is freely available and can accelerate such efforts in a variety of complex genetic disorders, as it can probe the cell type-specific effects of multiple non-coding SNPs across the global architecture of cell signalling and gene regulatory networks (https://github.com/korcsmarosgroup/iSNP).

In summary, by embedding and propagating signals from non-coding SNPs across downstream cell signalling and gene regulatory networks, our integrated network analysis provided plausible mechanisms connecting disease-associated genotype with pathogenic pathways, which were patient- and/or cell type-specific, and also linked to clinically relevant phenotypes in two distinct but related chronic IMIDs. In doing so, this work lays the foundations for addressing the challenges of “missing heritability” ^7,8^ and “missing regulation” ^11^, as well as genetic epistasis, that plagues complex disease genetics and which together pose a major barrier for the development of precision medicine strategies. Overall, this work underscores the importance of using systems genomics approaches to more accurately model and predict the holistic impact of non-coding risk variants in complex disease pathogenesis.

Whilst this systems genomics analysis led to plausible predictions for the impact of non-coding SNPs, we acknowledge the limitations of the presented study. We have not yet integrated non-regulatory SNPs such as coding risk-variants or considered the epigenetic architecture of the genome which has known effects in UC, CD, and other chronic IMIDs ^68^. These additional information layers could potentially hone our model. The lack of availability of a well-powered single-cell transcriptomics dataset in UC patients limited our ability to generate cell type-specific predictions for the various UC patient clusters we identified through our pipeline. Nevertheless, using bulk transcriptomics data, we demonstrated that UC SNP-propagated regulatory and cell signalling networks overlapped with UC inflammatory gene expression signatures from multiple independent studies, which further validated the power of our network propagation model to capture disease relevant genotype-to-phenotype mechanisms. Furthermore, there was insufficient clinical metadata in our patient cohort to correlate the gene regulatory network patient clusters with salient clinical features such as disease activity and phenotype, which could have provided further translational insights to our findings. However, large-scale biobanks for CD and UC (and other IMIDs) are currently being assembled which should provide a wealth of clinical metadata alongside various omics data that can be utilised for this purpose in the future ^69^ such as the IBD BioResource in the UK ^70^.

Tailoring therapeutic strategies to patient-specific pathogenic models will be critical for achieving precision medicine and improving patient outcomes in complex genetic diseases such as CD and UC. We believe that this study and workflow represents an important advancement towards this goal and can be readily applied to unravel the genetic underpinning of other chronic IMIDs.

### Availability of data and material

https://github.com/korcsmarosgroup/iSNP

## Data Availability

The patient genomic data is available: https://www.ibd-leuven.com/ibd-biobank
The code is available: https://github.com/korcsmarosgroup/iSNP
The network visualizations are available: https://www.ndexbio.org/#/networkset/5618007c-5257-11ee-aa50-005056ae23aa?accesskey=3de7d45d693d83e2fd96029b0d856f27aaa6e82337193346bbfc4621a59cbd60

## Acknowledgements

DM and AB were funded by a European Research Council Starting Grant (336159).

JPT is supported by the Chain Florey Clinical PhD Fellowship jointly funded by the National Institute for Health Research (NIHR) Imperial Biomedical Research Centre (BRC) and the UKRI Medical Research Council (MRC) Laboratory of Medical Sciences (LMS).

DM acknowledges financial support from Imperial College London through an Imperial College Research Fellowship grant award.

The work of DM, PS, MSB, LJH, SC and TK were supported by the BBSRC Gut Microbes and Health Institute Strategic Programme BB/R012490/1 and its constituent projects BBS/E/F/000PR10353 and BBS/E/F/000PR10355.

MP was supported by the UKRI Biotechnological and Biological Sciences Research Council (BBSRC) funded by Norwich Research Park Biosciences Doctoral Training Partnership (grant numbers BB/M011216/1 and BB/S50743X/1).

DM, PS, TK, SRC were also supported by a BBSRC Core Strategic Programme Grant for Genomes to Food Security (BB/CSP1720/1) and its constituent work packages, BBS/E/T/000PR9819 and BBS/E/T/000PR9817.

SRC, TK and DM were supported in part by the UK Biotechnology and Biological Sciences Research Council (BBSRC) under grant numbers BB/J004529/1, BB/R012490/1 and BBS/E/F000PR10355.

BV is funded by the Clinical Research Fund (KOOR), University Hospitals, Leuven, Belgium and the Research Council KU Leuven.

MM was founded through the UKRI BBSRC NRP DTP as a National Productivity Investment Fund CASE Award in collaboration with BenevolentAI (BB/S50743X/1)

TK and SRC were also supported by the UKRI BBSRC Institute Strategic Programme Food Microbiome and Health BB/X011054/1 and its constituent project BBS/E/F/000PR13631.

NP is supported by the Wellcome Trust (WT101159), Crohn’s and Colitis UK, and the NIHR Imperial Biomedical Research Centre (BRC)

TK and NP were supported by the NIHR Imperial Biomedical Research Centre (BRC). The views expressed are those of the authors and not necessarily those of the NIHR or the UK Department of Health and Social Care.

## Author contributions

DM, JBW and TK designed the updated iSNP workflow. JPT, JBW, DM and TK wrote the manuscript. BV provided the initial SNP data. JBW, DM, PS, MM, DF, LC, BB and MM developed and automated the workflow. DM and MP carried out network analysis and enrichment analysis. BAD, AZ, DC, JPT were involved in data analysis and interpretation. SC was involved in developing the iSNP workflow, data interpretation, and contributed to writing the manuscript. JBW, JPT, BV, NP provided clinical insight and/or clinical data analysis, and all contributed to writing the manuscript. DM and WXK carried out the transcriptomic validation. AB supervised the work of DM, BAD and AZ, and contributed to writing the manuscript. All the authors read and approved the final version of the manuscript.

## Materials and Methods

### Source of SNP data

Existing Immunochip data at the Leuven IBD unit were used for the current analysis (1,695 Crohn’s disease and 941 ulcerative colitis). All patients included in the analysis had given written consent to participate in the Institutional Review Board approved IBD Biobank (B322201213950/S53684).

Demographics data of the patients is available in Table S1.

### SNP annotation

For filtering the SNPs and annotating the affected regulatory interactions, we used the same methods as we have earlier described in Brooks-Warburton *et al.* ^17^. Briefly, we filtered the SNPs for ulcerative colitis and Crohn’s disease specific SNPs based on landmark genome wide association studies ^4,71^, and mapped these SNPs to the Immunochip data of our cohort. The disease-specific SNPs were annotated based on whether they were present in enhancers, promoters or miRNA-target sites and whether the SNPs altered these regulatory interactions. A gene and its translated protein was termed as “SNP-affected” if the SNP created or knocked out a transcription factor binding site or miRNA target site.

### Network propagation algorithm with random seed control

To build up the networks we used the seeds from genes with modified transcription factor-binding sites or miRNA target sites and used a directed HotNet2 algorithm ^72^ from the implementation of Huang et al ^41^. In brief, the algorithm creates a kernel from the degree weighted adjacency matrix with an alpha retain parameter, which describes how much heat is retained at each node. (Equation 1)

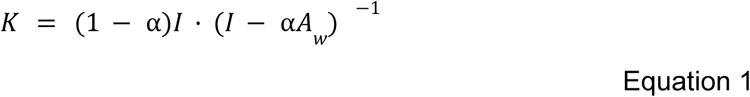

Where K is the resulted kernel matrix, *α* is the retain parameter, I is the identity matrix and A_w_ is the weighted adjacency matrix, where each edge is weighted according to the reciprocate of the out-degree of the source node.

*α* retain parameter was calculated using the following equation:

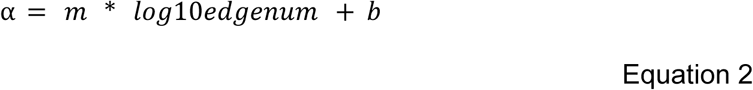

Here edgenum is the number of edges in the graph m and b are parameters, which were used as proposed by Huang et al m=-0.02935302 and b=0.74842057.

A unit heat was propagated through the K kernel matrix based on Equation 3.

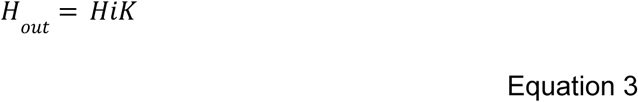

Here, the H_i_ is a diagonal matrix with unit (1) value for heat at each SNP-affected protein, and K is the kernel. Note if a patient has multiple SNPs then the network will have a higher amount of heat to distribute. If the heat reached a transcription factor, then it was propagated one step forward using a transcription factor-target gene network.

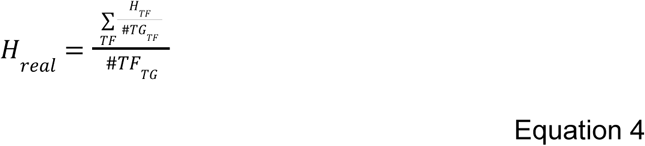

Here H_TF_ is the heat which reaches a transcription factor TF according to Equation 3. #TG_TF_ is the number of targets of a transcription factor TF. We sum the heat which proportionally affects each TG from each TF. #TF_TG_ is the number of transcription factors which affect target gene TG.

To avoid citation bias of the network resources, random seed controls were used. Here for each patient exactly the same number of seed genes were used for initial analysis. After 1000 repetitions from these random controls, the propagated heat resulted in a distribution. Z score was calculated from each distribution Equation 5.

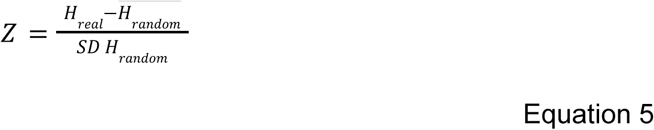

Where H_real_ is the heat of each node. H_random_ is the heat vector distribution using the random control. SD is the standard deviation and 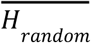 is the average of the random measures. If Z was > 2 then the protein was significantly affected. The Z score was similarly calculated in the case of transcription factor-target gene analysis.

The heat propagation by the kernel is analogous to a random walk with a restart method and represents a steady state (citation). Meanwhile, the regulatory one step heat propagation represents one step transcriptional response.

The analytical code is accessible in the GitHub repository of the Korcsaroslab at https://github.com/korcsmarosgroup/iSNP.

### Network resources

Signalling interactions data were obtained from the OmniPath database in October 2021 ^22,73^. The directed network giant component was used. It consists of 6954 protein nodes and 51,792 edges. For transcription factor-target gene interactions, we used the DoRothEA database with confidence scores A, B and C ^23^. This network contained 15,760 nodes and 90,145 edges.

### Overrepresentation analysis

For Gene Ontology and Reactome pathway overrepresentation analysis, we used the clusterProfiler (4.6.0) ^74^ package in the R environment (4.2.2) ^75^. In the case of Reactome with ReactomePA package (1.42.0) ^76^. The background used was OmniPath in the case of protein-protein interaction network analysis and DoRothEA in the case of transcription factor-target gene analysis. The enrichment plots were visualised using the enrichplot package (1.18.3). Identifier mapping was done using the org.Hs.eg.db (3.16.0) through AnnotationDBI (1.60.0).

### Visualisation and network analysis

Cytoscape 3.8 was used for visualisation ^77^. In the case of the SNP-propagated signalling networks the network modules were created using the clusterMaker app ^78^ GLay clustering, which is an implementation of the Grivan-Newman algorithm ^79^. In short, this algorithm removes the highest betweenness edges from the network till the network falls into multiple components. These components are considered as the new network modules. The UC regulatory network was modularised similarly. In the CD network, the nodes were moved by their involvement in various patient clusters by hand. If a gene was involved in a patient cluster more than 50% of the time, then it moved together with other genes from the same cluster. This resulted in a network which represented the patient specific cluster better. The networks are uploaded to the NDEX network resource (https://www.ndexbio.org/#/networkset/5618007c-5257-11ee-aa50-005056ae23aa?accesskey=3de7d45d693d83e2fd96029b0d856f27aaa6e82337193346bbfc4621a59cbd60).

### Patient-specific clustering

The patients were clustered using their SNP-propagated regulatory networks as a binary matrix as an input. First a Jaccard distance was calculated ^80^ using SciPy pdist function (v1.7.3) ^81^. The k-medoid clustering was used for clustering the patients due to the k-medoid giving better results in using binary features compared to k-means because the mode is better at summarising categorical features than the average ^82^ The used package was sklearn_extra, version: 0.2.0. Hierarchical clustering only produced small clusters due to the presence of outlier patients in both diseases. The optimal number of clusters was determined using the silhouette score ^83^ and the elbow method ^84^ implemented in the scikit-learn package (1.0.2) (Supplementary Figure 3) ^85^.

### Single-cell RNA-seq comparison

For single-cell RNA-seq validation, the Kong *et al* dataset was used for CD ^31^. The data set was downloaded from the Broad Institute Single Cell portal ^86^. The downloaded data was analysed by Seurat version 4.3.1 ^87^ in an R environment. We considered only cells which were expressing at least 200 genes and had a mitochondrial gene count below 5% and genes which were present in at least 3 cells. The data was then log normalised using Seurat’s NormalizeData function using standard parameters and scaled using Seurat’s ScaleData function using standard parameters. From the resultant Seurat object, using the FindMarkers function comparing cells originating from healthy and non-inflamed Crohn’s disease patients, differentially expressed genes (DEGs) were identified (adj p value<0.1, |log2FC| >0.5). These genes were used as the input for overrepresentation analysis comparing the cluster-specific genes in Crohn’s disease patients.

We ran a similar analysis using the recently published Krzak et al Crohn’s disease dataset as well. We downloaded the data from Zenodo https://zenodo.org/records/10528153l. We used the standard scanpy (1.9.6) pipeline in python (3.8). Filtering was performed as per Krzak et al, with the following excluded: cells with fewer than 100 genes expressed at ≥ 1 or mitochondrial count > 50, and genes expressed (>=1 count) in 5 or fewer cells. The counts were then normalised to 1e5, scaled and log transformed. We used the discovery cohort and the more granular cell type classification. We used python DEseq2 (version 0.4.10) pseudo bulk analysis comparing Crohn’s disease patients with healthy controls. Only cell types with a minimum of 10 cells in each category and minimum 15 cells in total were used and genes were filtered to have minimum 1000 counts. We defined differentially expressed genes from the pseudo bulk results as Benjamini-Hochberg adjusted p value <0.05 and absolute FC > 0.5. Note that the analysis of the two datasets uses slightly different pipelines to test the robustness of the outcome.

We performed a similar analysis on the Smillie *et al* dataset, which was at the time, the most comprehensive single-cell RNA-seq dataset in UC ^37^. However, we used slightly different filtering parameters due to the different quality of the dataset. Samples with less than 15% of mitochondrial transcript count and more than 200 genes, but less than 5000 expressed genes were kept. Following this, we conducted the analysis with the exact same parameters as those applied to the Kong *et al* dataset.

### Pseudobulk analysis for transcription factor activity

Both the Kong et al and the Krzak et al datasets were used to validate the activity of transcription factors predicted to be perturbed in the iSNP pipeline. For the Kong et al analysis we used a Seurat (4.3.1) pseudobulk pipeline comparing healthy controls with non-inflamed Crohn’s disease patients. We used the DeSeq2 (version 1.42.1) for differentially expression analysis and kept all fold changes. After identifying differentially expressed games we mapped the HGNC gene names to uniprot swissprot IDs using the ENSEMBL Biomart package (version 2.58.2), we ran a univariate linear model determining the activity of transcription factors using the decoupler package (version 2.9.7).

In the case of Krzak et al we ran the analysis in python because the data was available in scanpy h5at format. The gene names were mapped to uniprot swissprot IDs through the pybiomart package (version=0.2.0). Here we run the decoupler-py (version 1.7.0) tool from the pseudobulk outcome of the pydesq2 using the same DorothEA network as the input network as in the network propagation analysis and fitted a cell type-specific univariate linear model.

### Enrichment analysis in the UNIFI cohort

The UNIFI induction trial was a randomised placebo-controlled study investigating the efficacy and safety of ustekinumab for the treatment of moderate-to-severe ulcerative colitis^38^. Mucosal biopsies were collected at baseline from 364 patients who received ustekinumab (at 6mg/kg or 130mg intravenously), 186 patients who received placebo. Probe sets associated with multiple ENTREZ gene identifiers were discarded. Intensity data for probe sets mapped to the same ENTREZ gene identifier were summarised through their geometric mean. Differential gene expression analysis between responders (n=56) vs. non-responders (n=302) was performed in R ^75^ using the limma package ^88^. Response to therapy was assessed 8 weeks after induction; for this study, it was defined as mucosal healing, requiring endoscopic improvement (i.e., Mayo score ≼ 1) and histologic improvement (i.e., neutrophil infiltration in < 5% of crypts, no crypt destruction, and no erosions, ulcerations, or granulation tissue).

### Transcriptomics validation

As part of the validation analysis, DEGs were obtained from analysing transcriptomics data from IBD and healthy controls (Table S7). Using GEO2R ^89^, the samples were normalised using “limma” (3.46.0) ^88^. DEGs are defined by the following parameters: |logFC | >0.5 and adjusted p value< 0.05 by Benjamini-Hochberg method. The following studies were used for comparison in CD: Vancamelbeke et al, 2017 ^90^; Verstockt et al, 2019 ^91^; Pavlidis et al, 2021 ^92^; and the following studies in UC: Arijs et al, 2018 ^93^; Vancamelbeke et al, 2017 ^90^; Van der Goten et al, 2014 ^94^. The complete list of differentially expressed genes can be found in our

GitHub page: https://github.com/korcsmarosgroup/iSNP. An over-representation test was conducted to see whether the SNP affected genes, the genes in the SNP propagated signalling network or SNP propagated regulatory network were enriched in the differentially expressed genes using the clusterProfiler package ^74^.

## Supplementary Information

### Supplementary Notes

#### Supplementary Tables

Table S1: Patient demographics

Table S2: Non-coding single Nucleotide Polymorphisms (SNPs), associated transcription factors/miRNAs, and their SNP-affected target genes in Crohn’s disease and ulcerative colitis

Table S3: Reactome overrepresentation analysis in various network modules

Table S4: Functional overrepresentations of Reactome pathways in the commonly perturbed genes/proteins

Table S5: Transcriptomic validation of the network propagation models Table S6: Single-cell RNAseq signatures and cluster enrichment results

#### Supplementary Figures

**Figure S1.**
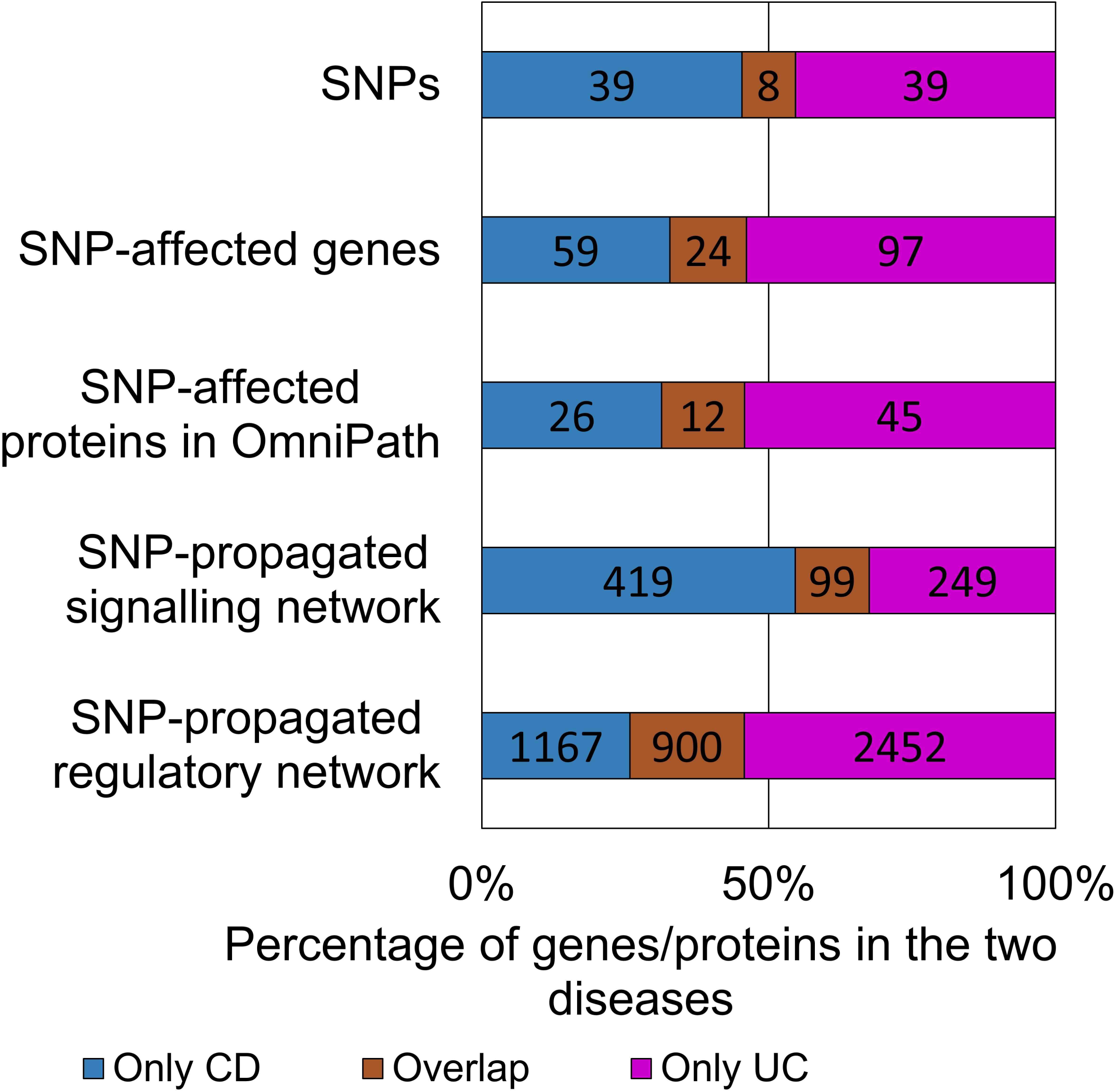
Number of features per propagation steps and disease. The number of SNPs in the cohort of CD and UC patients, along with the corresponding number of SNP-affected genes/proteins and SNP-propagated genes in the downstream signalling and regulatory networks.

**Figure S2.**
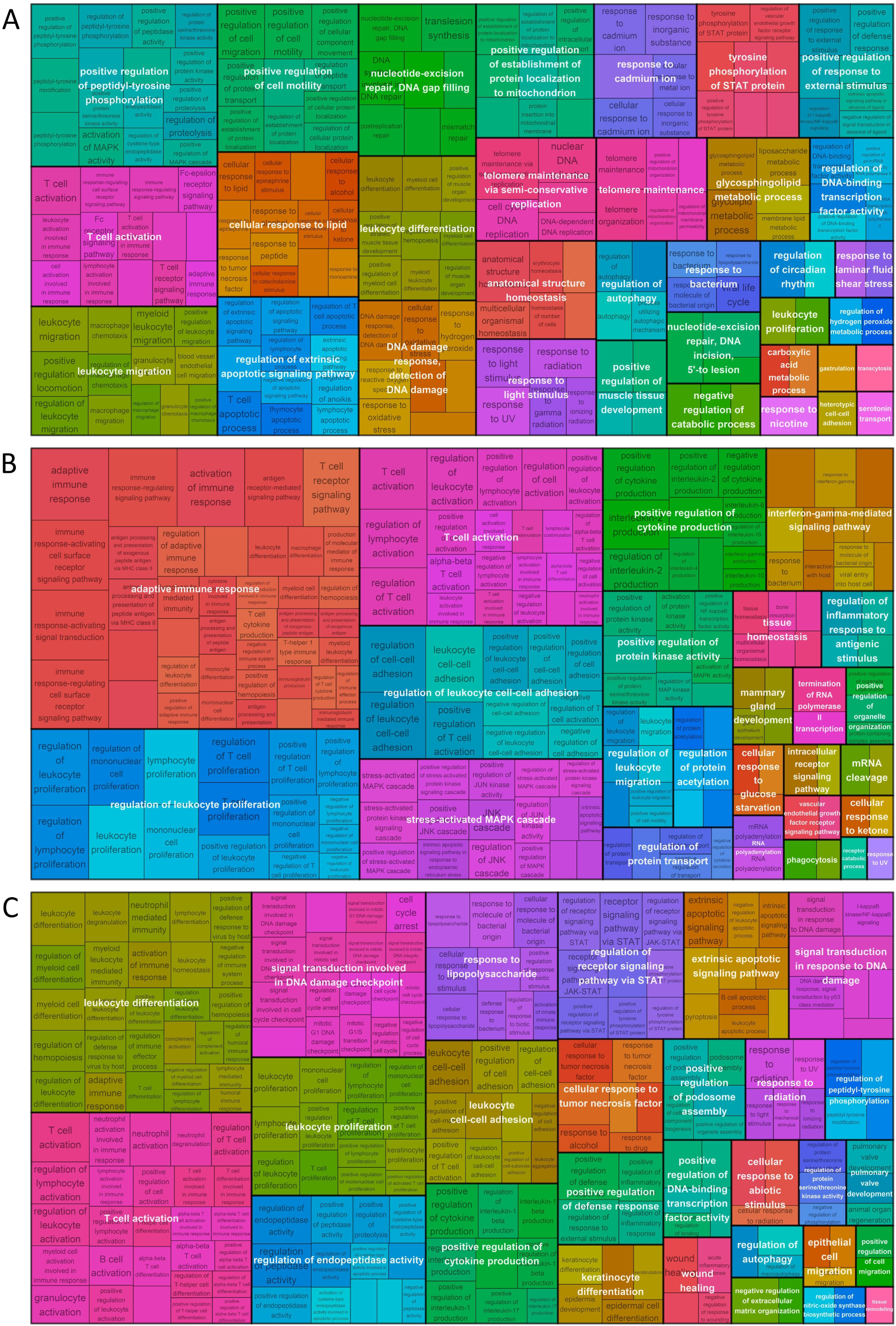
Tree map plots of the overrepresented Gene Ontology Biological Processes in the various SNP-propagated networks. A) SNP-propagated signalling network of Crohn’s disease patients. B) SNP-propagated signalling network of ulcerative colitis patients. C) SNP-propagated gene regulatory network of Crohn’s disease patients. The SNP-propagated gene regulatory network of ulcerative colitis patients had no enriched Gene Ontology Biological Processes.

**Figure S3.**
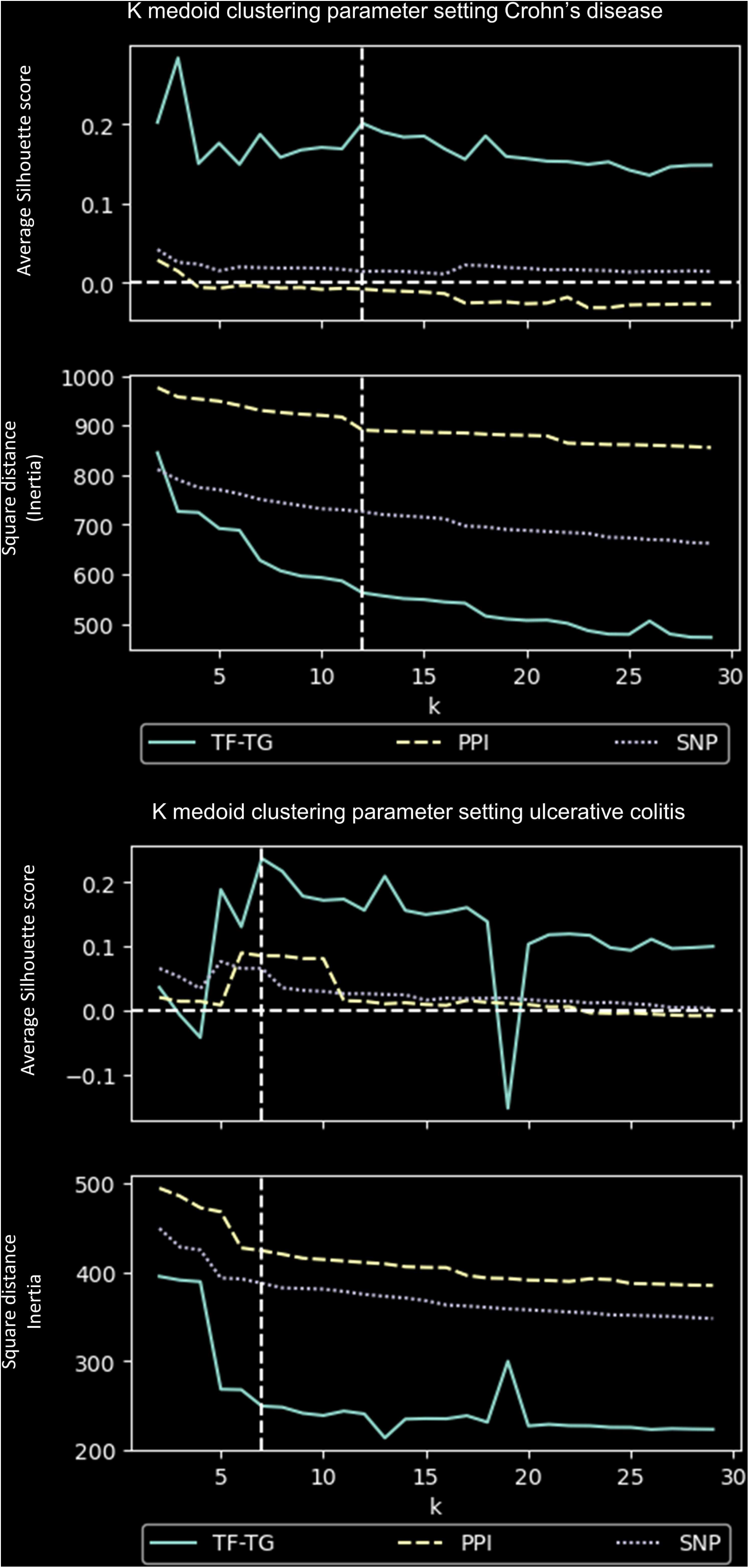
Determining the number of patient clusters in Crohn’s disease and ulcerative colitis using k medoid clustering. A) Crohn’s disease B) Ulcerative colitis. Each figure contains the SNP-affected genes, the PPI network and the regulatory networks as input for clustering. The regulatory networks have the highest average silhouette score and the steepest decline in inertia.

**Figure S4.**
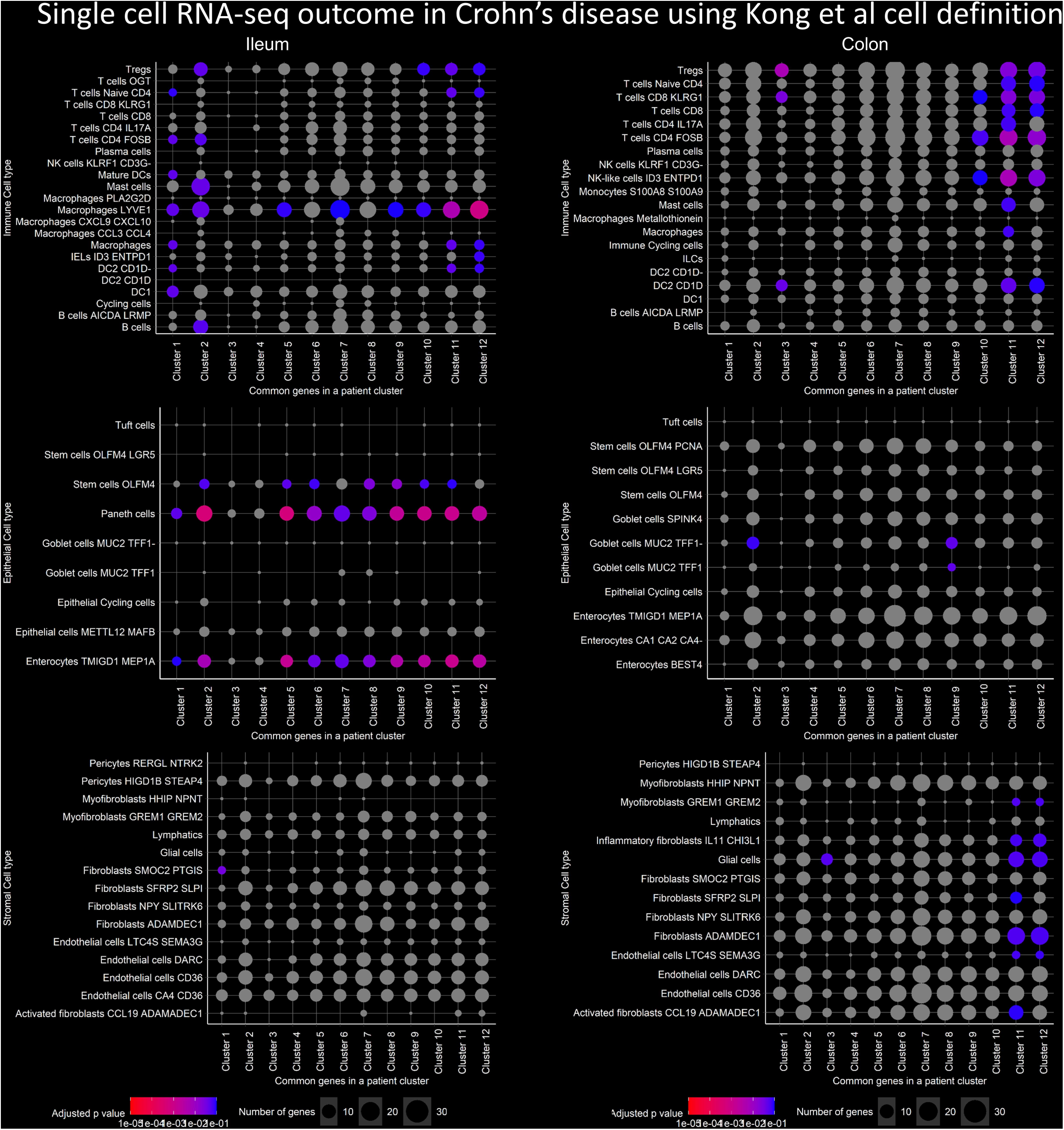
Overrepresentation analysis differentially expressed genes per cell type compared to patient clusters representing genes. Differentially expressed genes were calculated comparing healthy and uninflamed conditions (|FC| >0.5 and Benjamini Hochberg corrected p value <0.1). A gene was assigned as representing a SNP-propagated gene regulatory network cluster if the gene was perturbed in at least in 50% of the patients within that cluster.

**Figure S5.**
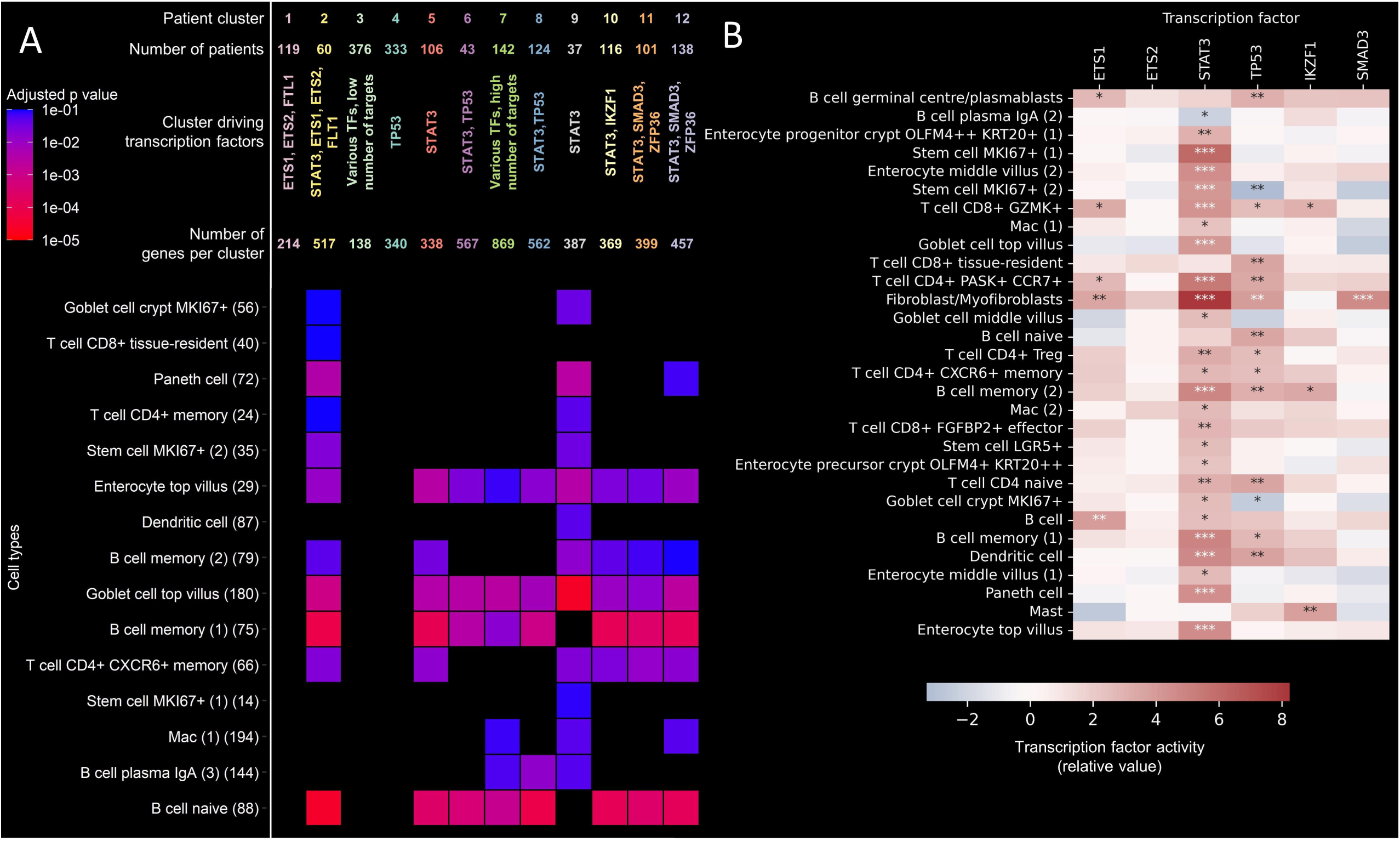
Validation on Krzak et al dataset. A) Differentially expressed genes are overrepresented in the various patient clusters. Figure legend is directly comparable and same as Figure 4. B) Transcription factor activity is cell specific in various cell types. Benjamini Hochberg adjusted p value * <0.05 **<0.01, ***<0.001, fitted slope of univariate linear model.

**Figure S6.**
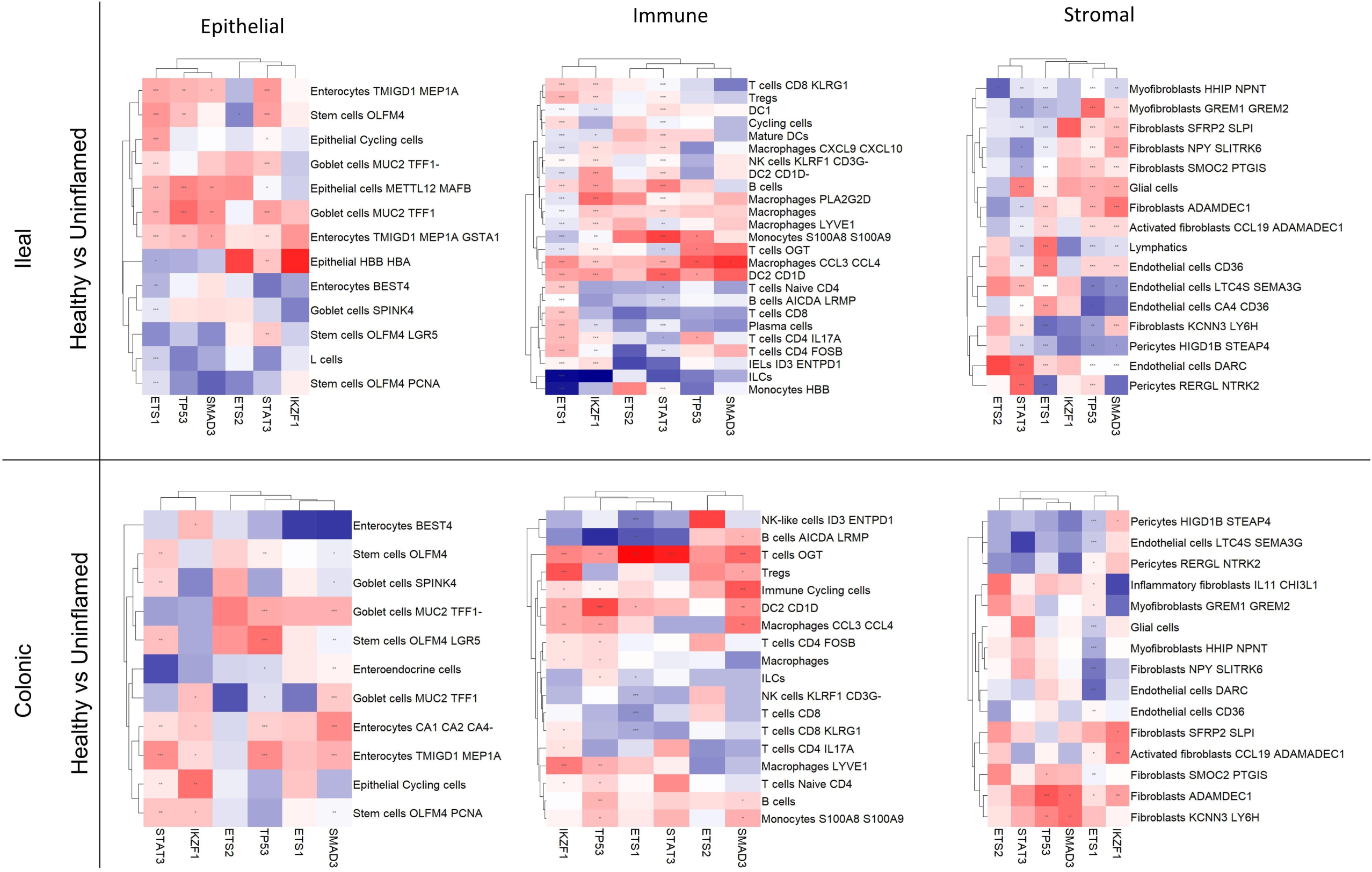
Regulatory analysis of transcription factors in Crohn’s disease Smillie et al dataset. Each subfigure represents a subset of cells in the Kong et al dataset. Red means active transcription factor in CD comparing the CD non-inflamed and healthy conditions meanwhile blue means inactive TFs. Benjamini Hochberg adjusted p value * <0.05 **<0.01, ***<0.001, fitted slope of univariate linear model.

**Figure S7.**
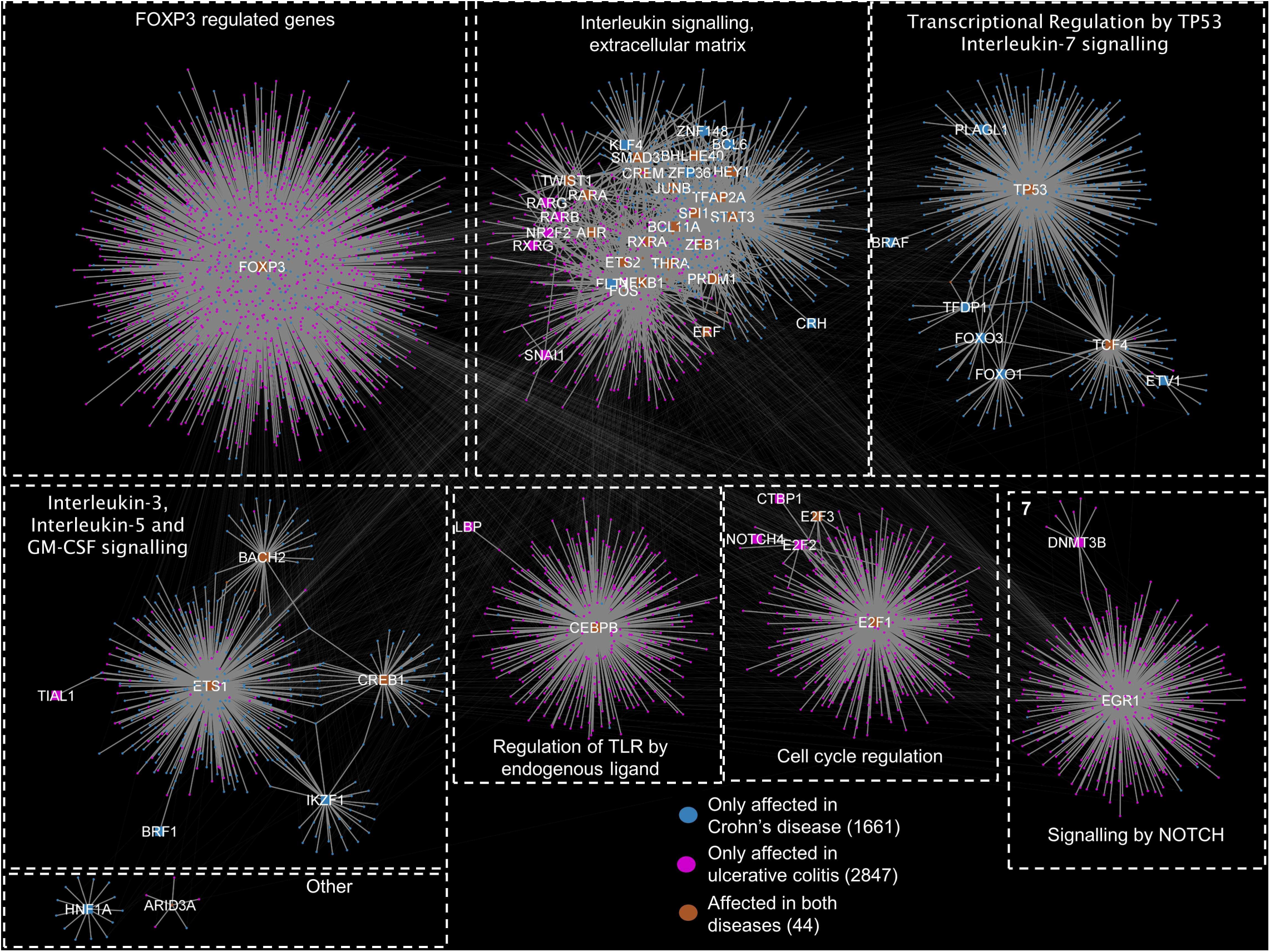
Combined SNP-propagated regulatory network for Crohn’s disease and ulcerative colitis. The network is modularised using the Girvan-Newmann algorithm in Cytoscape. Most modules are disease-specific. However, module 2 contains transcription factors and target genes that are present in both ulcerative colitis and Crohn’s disease patients.

