## Supplementary notes for "Decoding Non-coding SNPs: Systems Genomics Modelling Dissects the Heterogeneity of IBD"

**Supplemental Notes**

We found that non-coding SNPs propagate towards key biological pathways in cellular signalling networks that are well-established in IBD pathogenesis. In CD, disease-associated SNPs were predicted to impact proteins involved in apoptosis [[1,2]](https://sciwheel.com/work/citation?ids=11396703,6596843&pre=&pre=&suf=&suf=&sa=0,0) and autophagy [[3]](https://sciwheel.com/work/citation?ids=56856&pre=&suf=&sa=0) which are both known drivers of CD pathogenesis (Figure 2). Indeed, one of the first genotype-to-phenotype associations discovered in CD was autophagy following the GWAS identification of a coding variant (rs2241880, T300A) affecting the key autophagy gene *ATG16L1* [[4]](https://sciwheel.com/work/citation?ids=69922&pre=&suf=&sa=0). Subsequent mechanistic studies revealed that hypomorphic autophagy due to *ATG16L1* deficiency results in unresolved endoplasmic reticulum stress in Paneth cells and the development of spontaneous ileitis in mouse models [[5]](https://sciwheel.com/work/citation?ids=3584965&pre=&suf=&sa=0). Our findings indicate that two CD-associated non-coding SNPs (rs3792112 and rs3828309) located in transcription factor binding sites, converge on and perturb autophagy by impacting signalling processes involving ATG16L1 and two other autophagy-related proteins (CLTCL1 and CLTC). This finding demonstrates that autophagy signalling can be perturbed in a larger subset of CD patients than those having the *ATG16L1* coding risk variant alone, suggesting that dysfunctional autophagy has a widespread role in CD pathogenesis. Our study could aid the development of autophagy-targeting therapies for CD by providing a better understanding of the genetic mechanisms underpinning autophagy dysfunction in a subset of patients. Additionally, some reports suggest certain biologic therapies in CD, such as anti-TNF agents, suppress autophagy [[6]](https://sciwheel.com/work/citation?ids=3907753&pre=&suf=&sa=0). In patients with non-coding SNPs impacting autophagy (rs2241880, rs3792112, rs3828309), anti-TNF therapy may further enhance autophagy suppression which could potentially lead to reduced chance of therapeutic response. Whilst rs2241880 in isolation was not predictive of anti-TNF response [[7]](https://sciwheel.com/work/citation?ids=10209065&pre=&suf=&sa=0), our findings suggest that additional non-coding SNPs could be cumulatively predictive of therapeutic response.

In UC, non-coding SNPs were predicted to perturb proteins involved in established disease-associated pathogenic pathways such as NFκB [[8]](https://sciwheel.com/work/citation?ids=6799819&pre=&suf=&sa=0), IL1 [[9,10]](https://sciwheel.com/work/citation?ids=12703201,11988077&pre=&pre=&suf=&suf=&sa=0,0), IFNgamma [[11]](https://sciwheel.com/work/citation?ids=637501&pre=&suf=&sa=0) and IL17 [[12,13]](https://sciwheel.com/work/citation?ids=718866,425152&pre=&pre=&suf=&suf=&sa=0,0) signalling, I and MHC-II based antigen presentation [[14]](https://sciwheel.com/work/citation?ids=14956905&pre=&suf=&sa=0) (Figure 5). The most commonly affected proteins in the SNP-propagated signalling network of UC patients were functionally enriched for signalling pathways related to T cell receptor activation and interferon gamma (IFNɣ). Altogether these findings suggest a previously unrecognised genetic underpinning for major pro-inflammatory innate (IL1 signalling) and adaptive (Th1 and Th17 axes) immune pathways [[15]](https://sciwheel.com/work/citation?ids=13676934&pre=&suf=&sa=0), mediated by non-coding SNPs in UC patients. Whilst demonstrating the link between genotype and certain pathognomonic pathogenic pathways in IBD, the SNP-propagated signalling networks also unveiled less-established pathways that may be impacted by non-coding SNPs such as fatty acid metabolism in UC, and mast cell chemotaxis and DNA damage repair in CD. Based on these findings, these are likely to be important pathogenic pathways in IBD and require further experimental characterisation.

Evaluation of SNP-propagated gene regulatory networks also provided novel insights into the biological pathways disrupted by disease-associated SNPs at the gene regulatory layer. In the CD SNP-propagated gene regulatory network (Figure 3), several biological pathways implicated in IBD pathogenesis were enriched that were driven by key transcriptional regulators. These included, neutrophil degranulation [[16]](https://sciwheel.com/work/citation?ids=14957031&pre=&suf=&sa=0) (regulated by STAT3 and BRAF), integrin cell surface interactions [[17]](https://sciwheel.com/work/citation?ids=9409001&pre=&suf=&sa=0) (regulated by SMAD3 and ZFP36), IL7 signalling [[18]](https://sciwheel.com/work/citation?ids=6949873&pre=&suf=&sa=0) (regulated by multiple transcription factors), and MAPK signalling [[19]](https://sciwheel.com/work/citation?ids=719658&pre=&suf=&sa=0) (regulated by ETS1, ETS2, FLT1). Non-coding SNPs in UC were predicted to impact the downstream regulation of pathways also relevant in IBD pathogenesis such as retinoic acid signalling and bile acid metabolism [[20,21]](https://sciwheel.com/work/citation?ids=6612746,12562222&pre=&pre=&suf=&suf=&sa=0,0) (regulated by CEBP, SNAI1, and other transcription factors such as NR2F2, RXRA), Notch signalling [[22,23]](https://sciwheel.com/work/citation?ids=3180497,1346142&pre=&pre=&suf=&suf=&sa=0,0) (regulated by EGR1), and interleukin signalling (regulated by NFᴋB1) (Figure 6). These findings illustrate how small regulatory changes influenced by non-coding SNPs contribute to IBD pathogenesis through mostly distinct pathways at the gene regulatory layer in UC and CD.

In the UC SNP-propagated gene regulatory network, FOXP3 was perturbed in 71% (641/941) of UC patients in our cohort. FOXP3 is a key transcription factor for T regulatory (Treg) cell development [[24]](https://sciwheel.com/work/citation?ids=2879773&pre=&suf=&sa=0) and it also inhibits Th17 development depending on the presence of extracellular cues [[25]](https://sciwheel.com/work/citation?ids=2043640&pre=&suf=&sa=0) In active UC, there is a known imbalance between the pro-inflammatory Th17 and anti-inflammatory Treg axes, which favours mucosal inflammation [[26]](https://sciwheel.com/work/citation?ids=15013236&pre=&suf=&sa=0). Based on our findings, we posit that this may be driven by a transcriptional dysregulation of FOXP3 due to the presence of non-coding SNPs. We observed in our cohort that only patient clusters that had both NFkB1- and FOXP3-perturbed modules in their SNP-propagated gene regulatory networks were enriched for differentially expressed genes  in ustekinumab responders and non-responders from the UNIFI trial [[27]](https://sciwheel.com/work/citation?ids=7701283&pre=&suf=&sa=0). Ustekinumab is a major biologic therapy in UC which works by targeting the p40 subunit of IL12 and IL23 thereby inhibiting Th1 and Th17 pathways, respectively [[28]](https://sciwheel.com/work/citation?ids=86394&pre=&suf=&sa=0). We hypothesise that the NFkB1- and FOXP3-affected patient clusters have a lower chance of responding to ustekinumab therapy because their genetic background perturbs the genes which differentiate between treatment response and non-response. Whilst the underlying mechanisms are yet-to-be-characterised, non-coding SNPs that perturb NFkB1 may augment multiple pro-inflammatory pathways including those that are not specifically targeted by ustekinumab [[29]](https://sciwheel.com/work/citation?ids=5052194&pre=&suf=&sa=0). Coupled with a transcriptional dysregulation of FOXP3 limiting the anti-inflammatory effects of the Treg axis in these patients, this may result in an inadequate therapeutic effect of ustekinumab. This hypothesis needs further confirmation using patient cohorts where both genetic and therapeutic response data are available, followed by experimental validation.

SNP-propagated gene regulatory networks also stratified the cohort of CD and UC patients into separate patient clusters, implying significant disease heterogeneity in both forms of IBD at the gene regulatory level. These patient-specific clusters of SNP-propagated gene regulatory networks corresponded to cell type-specific differential gene expression patterns in CD. For instance, two SNP-propagated patient clusters regulated by ETS1 and ETS2 were enriched in several immune (e.g. macrophages, CD4 T cells, and dendritic cells) and epithelial cell types (e.g. enterocytes and Paneth cells) predominantly in the ileum, whilst two clusters regulated by STAT3, SMAD3, and ZFP36 mapped to various ileal and colonic immune, epithelial and stromal cell types. Corroborating these findings, recent work using *in vitro* models supports a critical role for ETS2 as a master regulator of pro-inflammatory responses in human macrophages which is perturbed by a pleiotropic risk variant (rs2836882) for multiple immune mediated disorders (including CD) located in the intergenic region on chr21q22 [[30]](https://sciwheel.com/work/citation?ids=14857386&pre=&suf=&sa=0). This specific risk variant was absent in our patient cohort, indicating that other non-coding SNPs can disrupt ETS2 function in a subset of CD patients not only in macrophages, but also in additional disease-relevant immune and epithelial cell types. This makes ETS2-regulated pathways an attractive therapeutic target in CD.

Finally, combined analysis and visualisation of both UC and CD SNP-propagated cell signalling networks highlighted pathways implicated in both IBD subtypes through shared and disease-specific SNP-propagated proteins. Functional analysis revealed that the distinct non-coding SNPs associated with UC and CD can affect the same intracellular processes such as T cell activity, apoptosis, and NFkB1 regulation, but through different disease-specific protein-protein interactions. Disease-specific signalling processes included autophagy in CD and CEBPB signalling in UC.  In contrast, little overlap was observed between the SNP-propagated gene regulatory networks of CD and UC patients (Figure 7C). For instance, FOXP3-target genes and STAT3-target genes were perturbed specifically in UC and CD, respectively. These findings further highlight that the impact of non-coding SNPs on downstream gene regulation are significantly different between CD and UC.  They also showcase the diverse putative routes originating from IBD-associated non-coding risk variants that traverse through downstream cell signalling and gene regulatory layers to converge on key pathogenic pathways in IBD. Consequently, we advocate combination therapies in IBD that target these key genotype-perturbed pathways from multiple angles. Preliminary insights regarding such an approach has been provided by the proof-of-concept VEGA trial which demonstrated that guselkumab (anti-IL-23 monoclonal antibody) and golimumab (anti-TNF monoclonal antibody) combination therapy was more effective than monotherapy in moderate-to-severe UC [[31]](https://sciwheel.com/work/citation?ids=14474440&pre=&suf=&sa=0).

**Supplementary Figures**
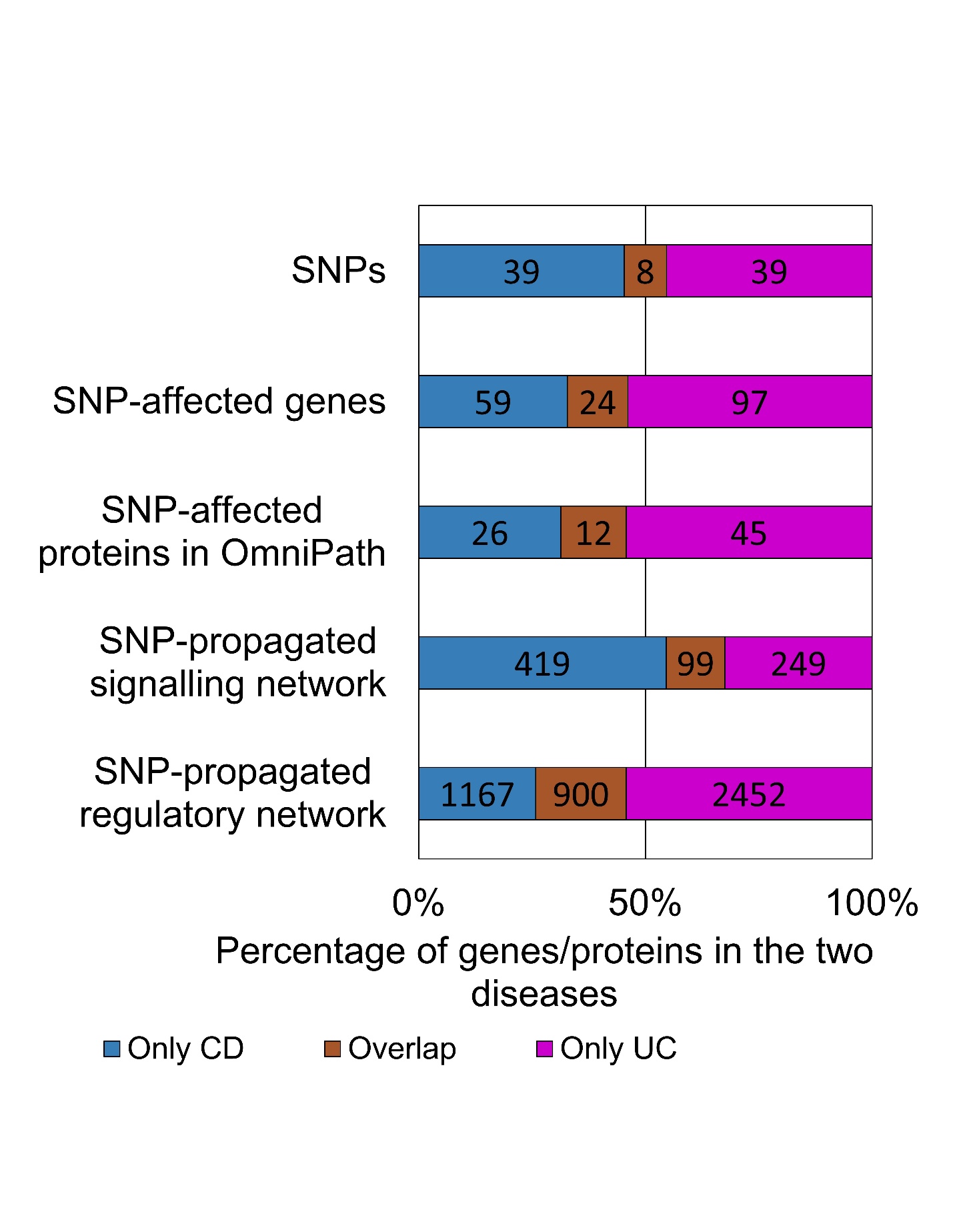
**Figure S1. Number of features per propagation steps and disease.** The number of SNPs in the cohort of CD and UC patients, along with the corresponding number of SNP-affected genes/proteins and SNP-propagated genes in the downstream signalling and regulatory networks.

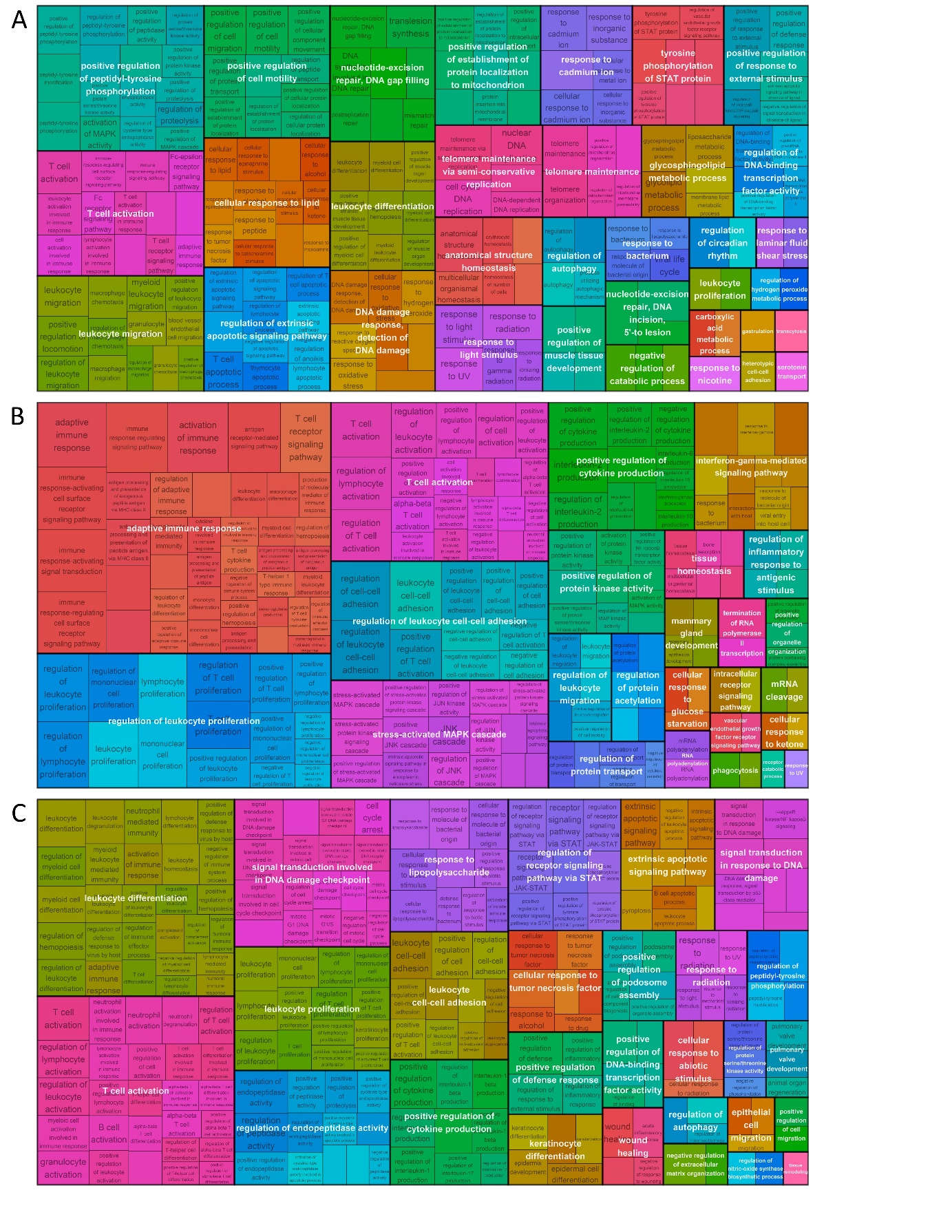

**Figure S2 Tree map plots of the overrepresented Gene Ontology Biological Processes in the various SNP-propagated networks.** A) SNP-propagated signalling network of Crohn’s disease patients. B) SNP-propagated signalling network of ulcerative colitis patients. C) SNP-propagated gene regulatory network of Crohn’s disease patients. The SNP-propagated gene regulatory network of ulcerative colitis patients had no enriched Gene Ontology Biological Processes.

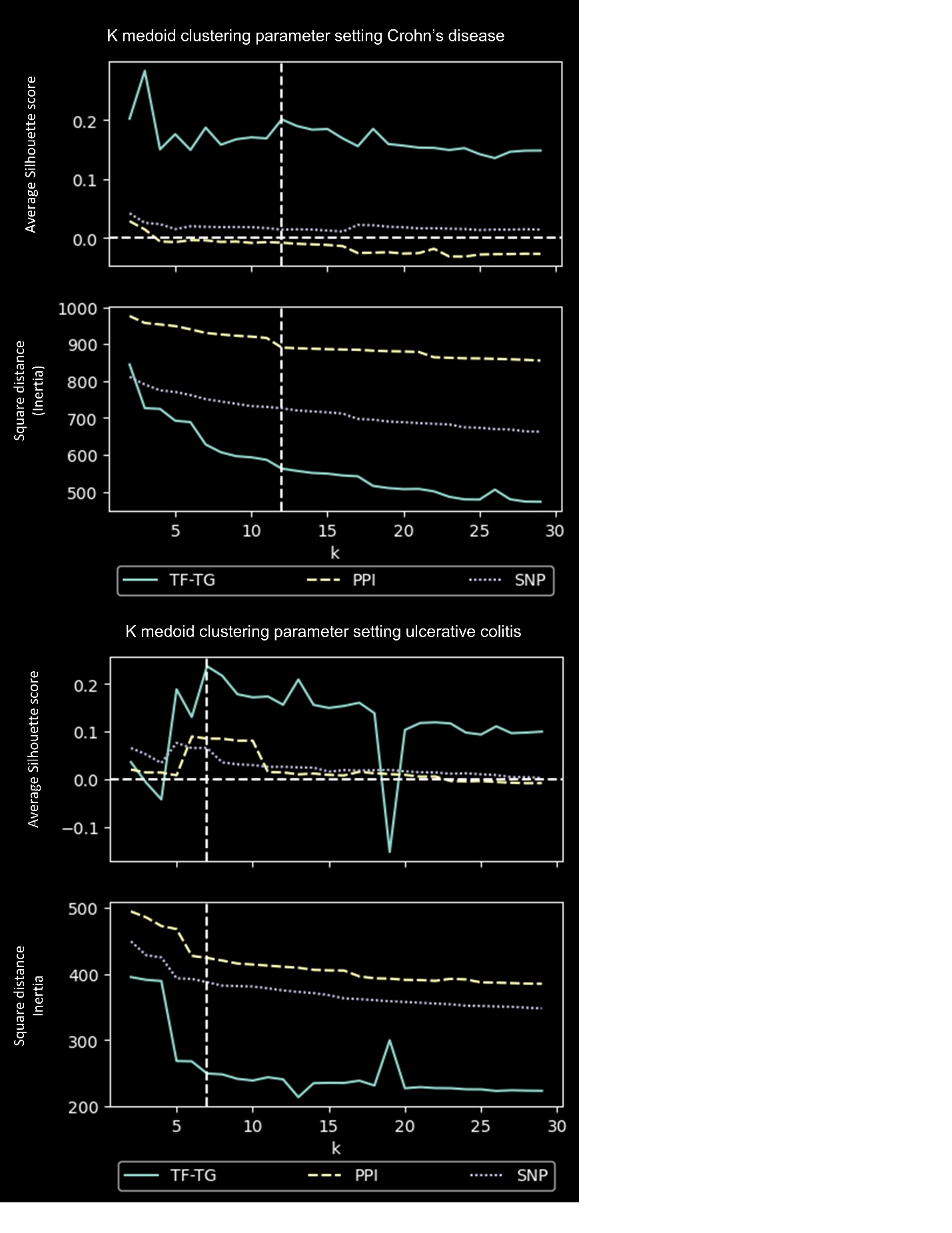

**Figure S3. Determining the number of patient clusters in Crohn’s disease and ulcerative colitis using k medoid clustering.** A) Crohn’s disease B) Ulcerative colitis. Each figure contains the SNP-affected genes, the PPI network and the regulatory networks as input for clustering. The regulatory networks have the highest average silhouette score and the steepest decline in inertia.

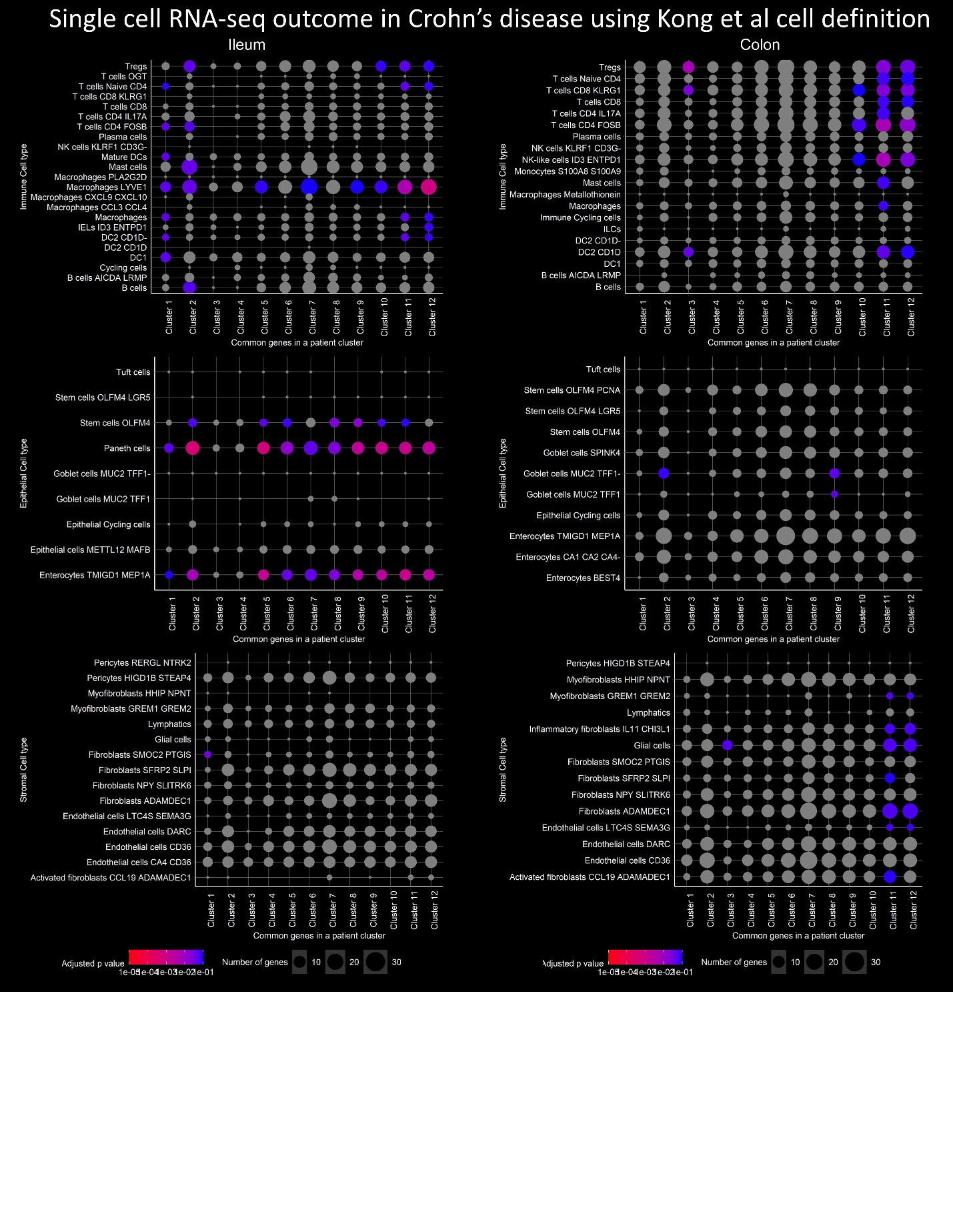

**Figure S4. Overrepresentation analysis differentially expressed genes per cell type compared to patient clusters representing genes.** Differentially expressed genes were calculated comparing healthy and uninflamed conditions (|FC| >0.5 and Benjamini Hochberg corrected p value <0.1). A gene was assigned as representing  a SNP-propagated gene regulatory network cluster if the gene was perturbed in at least in 50% of the patients within that cluster.

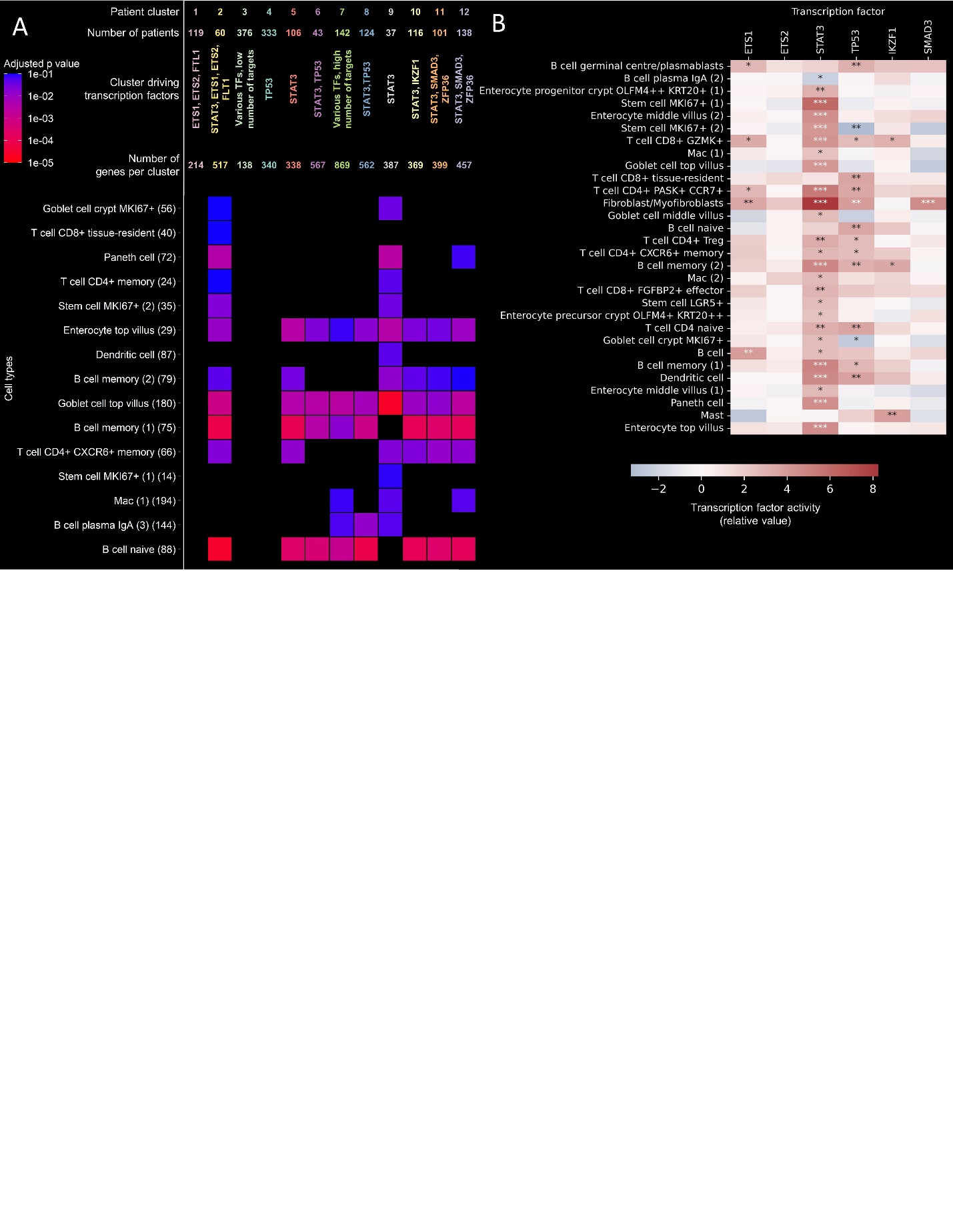
**Figure S5. Validation on Krzak et al dataset.** A) Differentially expressed genes are overrepresented in the various patient clusters. Figure legend is directly comparable and same as  Figure 4. B) Transcription factor activity is cell specific in various cell types.  Benjamini Hochberg adjusted p value * <0.05 **<0.01, ***<0.001, fitted slope of univariate linear model.

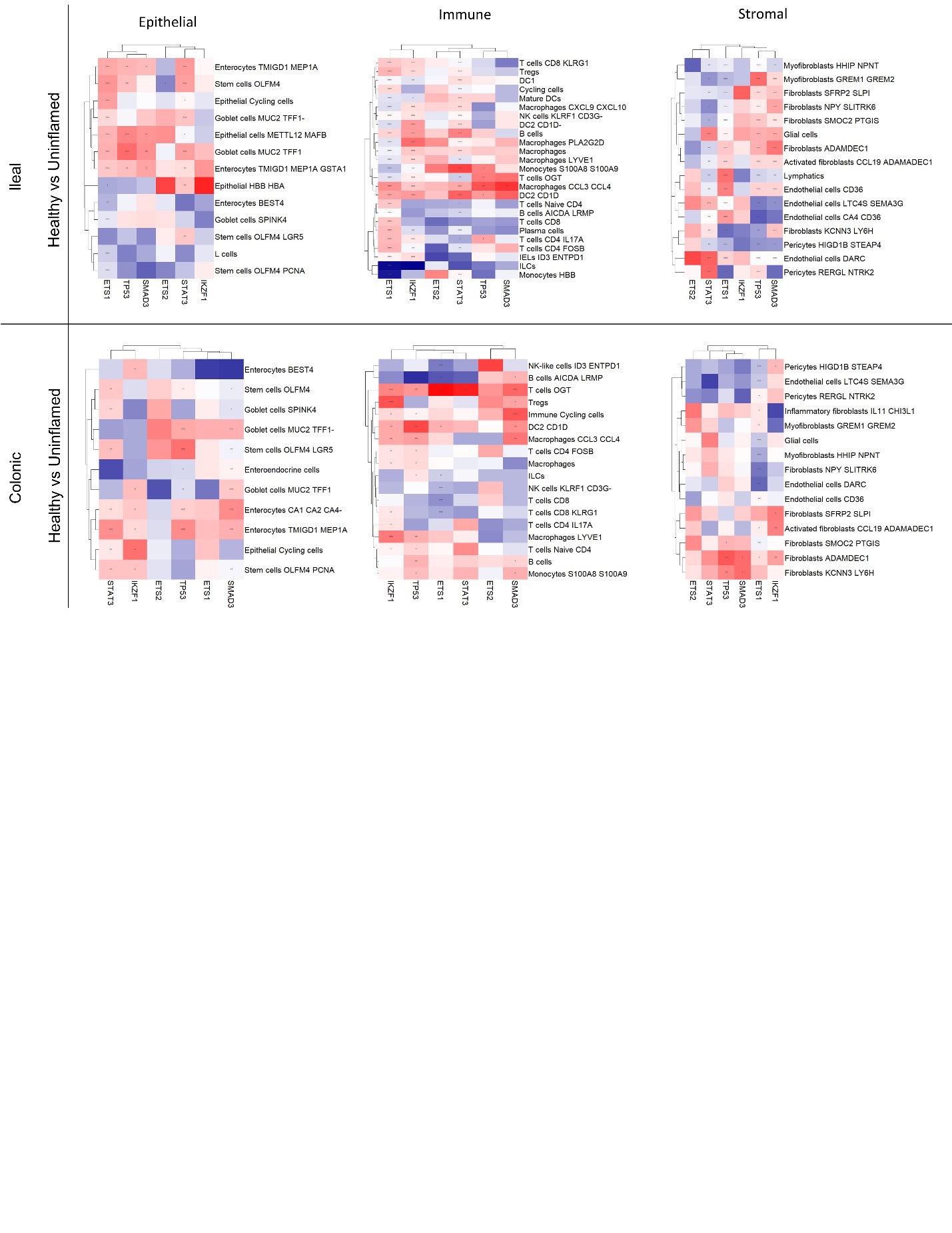

**Figure S6. Regulatory analysis of transcription factors in Crohn’s disease Smillie et al dataset.** Each subfigure represents a subset of cells in the Kong et al dataset. Red means active transcription factor in CD comparing the CD non-inflamed and healthy conditions meanwhile blue means inactive TFs. Benjamini Hochberg adjusted p value * <0.05 **<0.01, ***<0.001, fitted slope of univariate linear model.

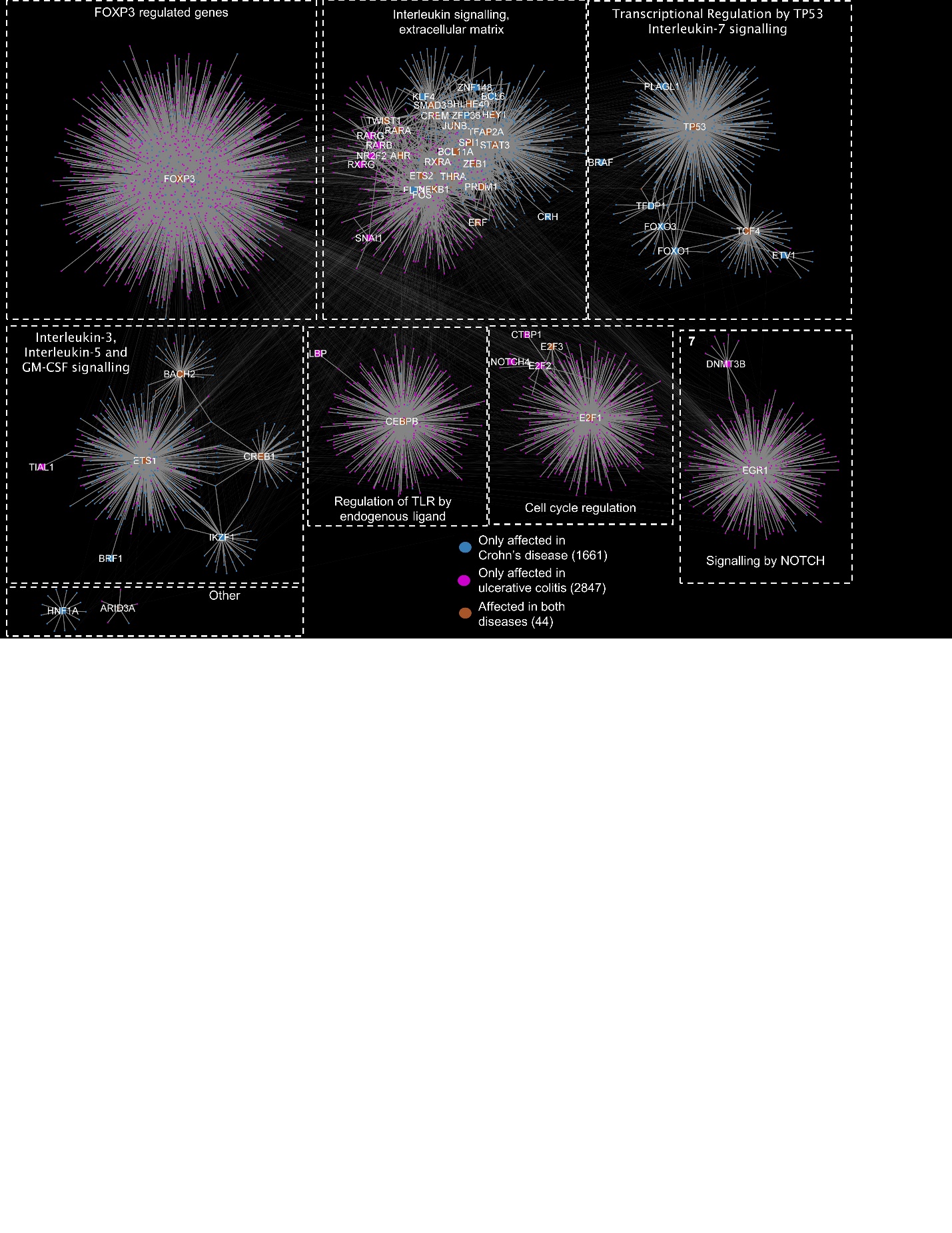

**Figure S7. Combined SNP-propagated regulatory network for Crohn’s disease and ulcerative colitis.** The network is modularised using the Girvan-Newmann algorithm in Cytoscape. Most modules are disease-specific. However, module 2 contains transcription factors and target genes that are present in both ulcerative colitis and Crohn’s disease patients.
